# Blood immune profiles reveal a CXCR3/CCR5 axis of dysregulation in early sepsis

**DOI:** 10.1101/2024.03.21.24304671

**Authors:** David Kealy, Julie Wilson, Tom Jaconelli, Karen Hogg, Rebecca Coop, Greg Forshaw, Neil Todd, David Yates, Nathalie Signoret

## Abstract

Sepsis is defined as a systemic inflammatory host response syndrome after serious microbial infection, which requires prompt treatment to lower the risk of complications and death. However, early sepsis recognition can be a challenge at presentation when patients show symptoms difficult to distinguish from other acute conditions.

We designed a pilot study to explore whether blood immune signatures could reveal early specific indicator profiles for patients meeting sepsis criteria upon admission at the hospital Emergency Department. We analysed blood samples from study-recruited sepsis-suspected patients (N=20) and of age-spanning healthy volunteers (N=12), using flow cytometry-based assays. 25 circulating inflammatory cytokines and chemokines (CCs) were measured from blood plasma, while freshly isolated unfixed blood leukocytes were immunophenotyped to ascertain major cell subsets representation and expression of activation markers, including chemokine receptors. We found that beside IL-6 and sCD14, blood levels of CXCL9 and CXCL10 (two ligands of CXCR3) show good separation between healthy controls and sepsis-suspected patients. The abundance of CD4^+^ T cells was significantly reduced while the expression of chemokine receptors was altered on monocytes, B and all T cells from patients. In particular, we report substantial losses of CCR5-expressing monocytes and CXCR3/CCR5 double positive T cells. Full dataset analysis and post-hoc subgrouping of patients according to their diagnosis on discharge (confirmed or unconfirmed sepsis), identified CXCR3/CCR5 double expression on T cells as a separating characteristic within the study. Overall, our observational study suggests a new CCR5 and CXCL9-10/CXCR3 axis of dysregulation in early sepsis.

## Introduction

Sepsis is a common life-threatening complication of bacterial infections, with sepsis-related mortality accounting for 20% of deaths worldwide in 2017^1^. Early administration of appropriate antibiotics is the mainstay of treatment and acting within an hour of patient admission lowers the risk of complications and death^2^. However, often at that time the causative agent of sepsis is unknown, and the clinical recognition of the condition can be challenging due to the heterogeneity of signs and symptoms^3, 4^. In fact, many patients meeting sepsis criteria upon admission in the emergency department (ED) are not assigned a diagnosis of sepsis by the time of discharge^5^. Being able to accurately diagnose sepsis cases within a few hours of their arrival in ED would allow for more targeted and effective antibiotic interventions and improve survival outcomes^6^. Since bloodstream infections leading to sepsis are rarely associated with high blood bacteraemia^7^ but do fundamentally alter inflammatory biomarkers in blood^8^, the emphasis has been on defining sets of bacterial and immune biomarkers that could be used as reliable and rapidly measured predictors of sepsis^4^. Among the multitude of biomarkers evaluated in the past two decades^9^, only procalcitonin (PCT), C-reactive protein (CRP) and the cytokine Interleukin 6 (IL-6) have shown some diagnostic and prognostic value. Although the levels of these three soluble inflammatory markers in blood can provide information on the severity of infection, their performance in differentiating bacterial from viral sepsis or non-infectious conditions with systemic inflammatory response syndrome (SIRS) is limited^10, 11^. It may be possible that widening the range of biomarkers studied would help define unique signatures distinguishing between these different conditions^12, 13, 14^.

The pathogenesis of sepsis is due to immune imbalance following an acute response to infection with the occurrence of two opposite host reactions, a pro-inflammatory SIRS and an immunosuppressive compensatory anti-inflammatory response syndrome (CARS)^15, 16^. This imbalance is not only associated with altered systemic levels of circulating inflammatory mediators such as cytokines and chemokines, but also with functional and phenotypic changes of innate and adaptive immune cells responding to these mediators^17, 18^. Therefore, identifying dominant cellular changes correlating with unique soluble biomarker profiles from blood samples may expose specific immune signatures that can discriminate sepsis from other forms of systemic inflammation. The dynamics of sepsis as a condition evolving from severe inflammation to SIRS and CARS means that identifying changes in inflammatory markers detected at one point in time to help sepsis diagnosis will present a challenge ^4^. However, identifying combinations of changed biomarkers could distinguish a sepsis signature at onset of clinical signs and symptoms^19^.

Cytokines, chemokines and their receptors are essential regulators of inflammation impacting on the recruitment, activation and function of leukocytes, but also account for imbalances in the inflammatory network leading to sepsis pathology^20, 21^. With regards to bacterial infection, a number of studies including ours have evidenced a direct effect of Gram^-^ or Gram^+^ bacteria cell wall components Lipopolysaccharide (LPS) or Lipoteichoic acid (LTA) on cytokines and chemokine production as well as receptor expression and cell activation^22, 23^. Despite this, sepsis-induced phenotypic changes affecting different subpopulations of circulating leukocytes remain poorly understood, particularly with regards to their expression of activation markers and chemokine receptors.

With these points in mind, we conducted a pilot study to identify potential immune signatures from blood of sepsis-suspected patients upon hospital admission compared to blood of healthy volunteers. Using flow cytometry-based multiplex assays, we measured plasma levels of circulating biomarkers that modulate inflammation and explored white blood cells parameters by performing immunophenotyping of live freshly isolated PBMCs. We assessed cell-surface markers that define specific immune cell subsets, their function and activation status, to identify parameters that significantly differ between sepsis-suspected patients and healthy controls. Using supervised data analysis, we identified the prevalent variables emerging from our datasets and their ability to discriminate between healthy controls and sepsis-suspected patients. We identified overlapping but distinct immune signatures between groups with differences in IL-6, sCD14, CXCL9 and CXCL10 levels, the expression of CCR5 on monocytes, the abundance of CD4^+^ T cells, plus the proportion of double positive CXCR3/CCR5 in CD4^+^ and CD8^+^ T cell subpopulations being the most prominent separating traits. Our study revealed that alterations in CCR5 expression and the CXCL9-10/CXCR3 axis could be useful early indicators of sepsis.

## Material and Methods

### Reagents and Antibodies

Tissue-culture reagents, all secondary antibodies, and conjugated-streptavidin were purchased from Thermo Fisher Scientific (Paisley, Renfrewshire, United Kingdom). Other reagents were from Sigma-Aldrich (Gillingham, Dorset, United Kingdom), unless stated otherwise. Fluorochrome-conjugated antibodies used (see Fig. S2) were purchased from BioLegend (San Diego, CA, USA) or Abcam (Cambridge, UK) with the exception of the anti-CCR5 antibody MC5^22, 24^, which was purified in house from hybridoma (gift from Prof. Matthias Mack University of Regensburg, Germany), and fluorescently conjugated using Invitrogen Alexa Fluor 488 or 647 antibody labelling kits (Thermo Fisher Scientific). In addition, we used a cell viability kit from Invitrogen Live/DEAD Fixable Near IR for 808 nm excitation (Thermo Fisher Scientific).

### Study design

The study was sponsored by the York & Scarborough Teaching Hospitals NHS Foundation Trust. It received ethical approval from Yorkshire & The Humber - Leeds West Research Ethics Committee (REC reference 19/YH/0394) for IRAS project ID: 269597. The clinical recruitment of patients presenting to the ED of The York Hospital with moderate to high risk of having sepsis, followed the National Institute of Clinical Excellence (NICE) Clinical Guideline 51^25^, which mandates that all these patients require blood sampling to confirm or refute the diagnosis. For inclusion to the study, patients had to present at least two abnormal physiological parameters identified in the NICE guideline for adults aged 18 and over in acute hospital setting presenting ‘Moderate to high risk’ ^25^ using the ‘Sepsis: Risk stratification tool’ ^26^ with symptoms or signs of chest infection, pneumonia and/or cellulitis to select for suspected infections of bacterial origin. After obtaining written informed consent, blood samples from recruited patients were collected for hospital microbiology tests with an extra 10 ml tube of blood drawn for the study. Each sepsis-suspected patient was assigned a unique study ID number, and blood samples were linked-anonymised for the biological analysis and reconciliation of results with clinical outcomes at the end of the study. Trial ID numbers were used to create an anonymised dataset compiling hospital results, microbiological outcomes and clinical information collected post-discharge, leading to a final diagnosis of confirmed or unconfirmed sepsis status (see Table S1).

A control cohort of healthy donors for age-matching representation was recruited from university healthy volunteers and consenting elderly healthy participants attending The York Hospital for elective orthopaedic surgery, with local ethical approvals. Healthy donors reported good general health and no recorded ongoing treatment for conditions that would impact their immunity.

### Blood sample collection and processing

For study recruited patients (N=20) and healthy controls (N=12), 9 ml of whole venous blood was collected in a S-Monovette K3 EDTA tube (Sarstedt, United Kingdom). All samples were linked-anonymised, stored at 4°C and transferred to the University of York research laboratory to be processed within a maximum of 6h post-collection. Whole blood samples were diluted 1:1 in PBS and added to a 50 ml Leucosep separation tube (Greiner Bio-One, United Kingdom) pre-equilibrated with 15 ml of Lymphoprep density gradient medium (StemCell Technologies) before centrifugation to separate the plasma fraction and recover the white blood cells layer (leukocytes) as previously described^27^. Plasma fractions were cryopreserved until biomarkers measure assays were performed, while leukocytes were used immediately for live cell immunophenotyping.

### Measurement of biomarkers in human plasma fractions

We used pre-designed multiplex flow cytometry beads-based assay panels (LEGENDplex^TM^ BioLegend, San Diego, CA, USA) to measure blood circulating levels of cytokines and chemokine from isolated plasma fractions. We used the Human Th cytokines panel 12-plex kit (IL-2, 4, 5, 6, 9, 10, 13, 17A, 17F, 21 and 22, IFNγ and TNFα) and the Human proinflammatory chemokine Panel 1 12-plex kit (CCL2, 3, 4, 11, 17, 20. CXCL1, 5, 8, 9, 10 and 11) plus a separate CCL5 1-plex kit, according to manufacturer instruction. Data were acquired on a CytoFLEX LX (Beckman Coulter, Indianapolis, IN, USA) flow cytometer and analysed using the LEGENDplex data analysis software to calculate concentrations that fall within the bounds of intra-assay generated standard curves (assay sensitivity 0.2-3.8 pg/ml depending on the analyte). Levels of serum released CD14 (sCD14) were assessed with a CD14 human ELISA kit (Invitrogen, Thermo Fisher scientific; assay sensitivity 6pg/ml).

### Immunophenotyping of live, freshly isolated Leukocytes

Isolated leukocytes (5-6 x 10^6^ cells) were resuspended in 1 ml of ice-cold FACS buffer (FB: PBS, 1% FCS, 0.05% sodium azide) and divided between 5 wells in a U bottom 96-well plate kept on ice, including two well for unstained cells and live/dead stain alone controls, plus wells for cell surface markers staining using either a Blood Cell Panel (BCP) for CD3, CD14, CD16, CD19, TLR2, CCR1, CCR2, CCR5 and CCR7 or a T Cell Activation Panel (TCAP) for CD4 CD8 (alpha chain), CD25, CD45RO, TLR2, CXCR3 and CCR5 (See Fig. S1). Cell were stained unfixed, with samples incubated on ice. Cells were first treated for 20 mins in 50µl of FB supplemented with 20μg/ml of human IgG to saturate F_C_ receptors before adding 50µl of the relevant staining panel and labelling for 1h on ice. Samples were washed three times 5 mins in 200µl of ice-cold FB before adding a 1:5000 dilution of Live/DEAD Fixable Near IR cell viability dye in ice cold PBS and incubate for further 20 mins on ice before washing in PBS alone, where required. Fluorescence Minus One (FMO) experimental controls generated using cells from healthy donors were used to set upper limits of background signal versus positive populations for each omitted fluorescent antibody from the multicolour panels BCP and TCAP (data not shown). Post-staining, samples were fixed O/N at 4°C in 250 µl of FACS FIX (FB with 1% methanol-free formaldehyde solution), washed and resuspended in 200 µl of FB before data acquisition by running 75µl per sample (90 secs at a flow rate of 30µl/min) on a CytoFLEX LX, results pre-analysed using the CytExpert Software (Beckman Coulter) and detailed multicolour analysis was performed with FCS Express De Novo Software (Dotmatics). Gating strategies used for data analysis are reported in Fig. S3). Results were expressed as the percentage of cells positive for the indicated cell-surface markers within the specified cell population(s) that data were gated on for each graph.

### Quantification, data integration and statistical methods

#### Univariate statistics

Data from each experiment were analysed with GraphPad Prism version 10 software using Mann Whitney or ANOVA with the indicated multiple comparison post-tests, where appropriate. Boxplots show 25th and 75th percentiles (boxes), medians (lines in boxes), with Tuckey whiskers and outliers, or minimum to maximum values whiskers for graphs showing all points. A measure of group separation achieved by individual variables was defined by:

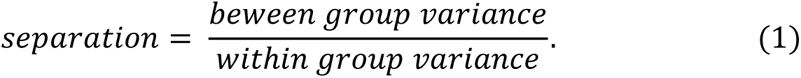

#### Multivariate statistics

Multivariate analyses were performed in the R programming environment^28^. Principle component analysis (PCA) was carried out using the function ‘prcomp’ in base R. As an unsupervised method, no information on group is used in PCA and any patterns related to group cannot therefore be forced. Partial least squares regression (PLSR) was performed using the R package ‘pls’ with the response variable encoded as 1.0 and 2.0 for Healthy and sepsis-suspected (Sepsis) respectively. Classification was achieved by assigning the class corresponding to the closest integer to the output response. Random Forest classification was performed using the R package ‘randomForest’. Due to the potential for overfitting in supervised analyses, we used leave-one-out (L-O-O) cross validation. For L-O-O classification, the class for each observation was predicted in turn from a model built without the data for that observation.

#### Spearman correlation

For all correlations between individual variables, Spearman’s rank-order correlation was used. Where there were missing values, correlations were calculated over all patients for which values were available. Heatmaps showing multiple correlations were created using the package ‘pheatmap’ in R.

## Results

### Participants characteristics and blood samples

A total of 20 patients admitted to The York Teaching Hospital ED with suspected sepsis (see Table S1), who matched inclusion criteria were enrolled; 45% were female and patients mean age was 70 (Fig. 1A). Blood samples analysed for the study were taken upon patient admission to ED, before hospital microbiological cultures were performed and a full clinical diagnosis of sepsis ascertained. Initial hospital white blood cell (WBC) counts for many patients were above the standard reference range^29^ (Fig 1.B). In parallel, 12 healthy controls were recruited from university volunteers and healthy older individuals attending The York Hospital for elective knee surgery for an age-matching healthy donor cohort, with a mean age of 51 and 58% female (Fig. 1A). For both groups, the number of cells recovered after gradient density isolation was within the accepted yield range^30^, with two exceptions in the sepsis-suspected group (Fig. 1C). These observations suggest that the dominant increase in WBC for sepsis-suspected patients relates to neutrophilia, a common consequence of bloodstream infection^31^.

**Figure 1:**
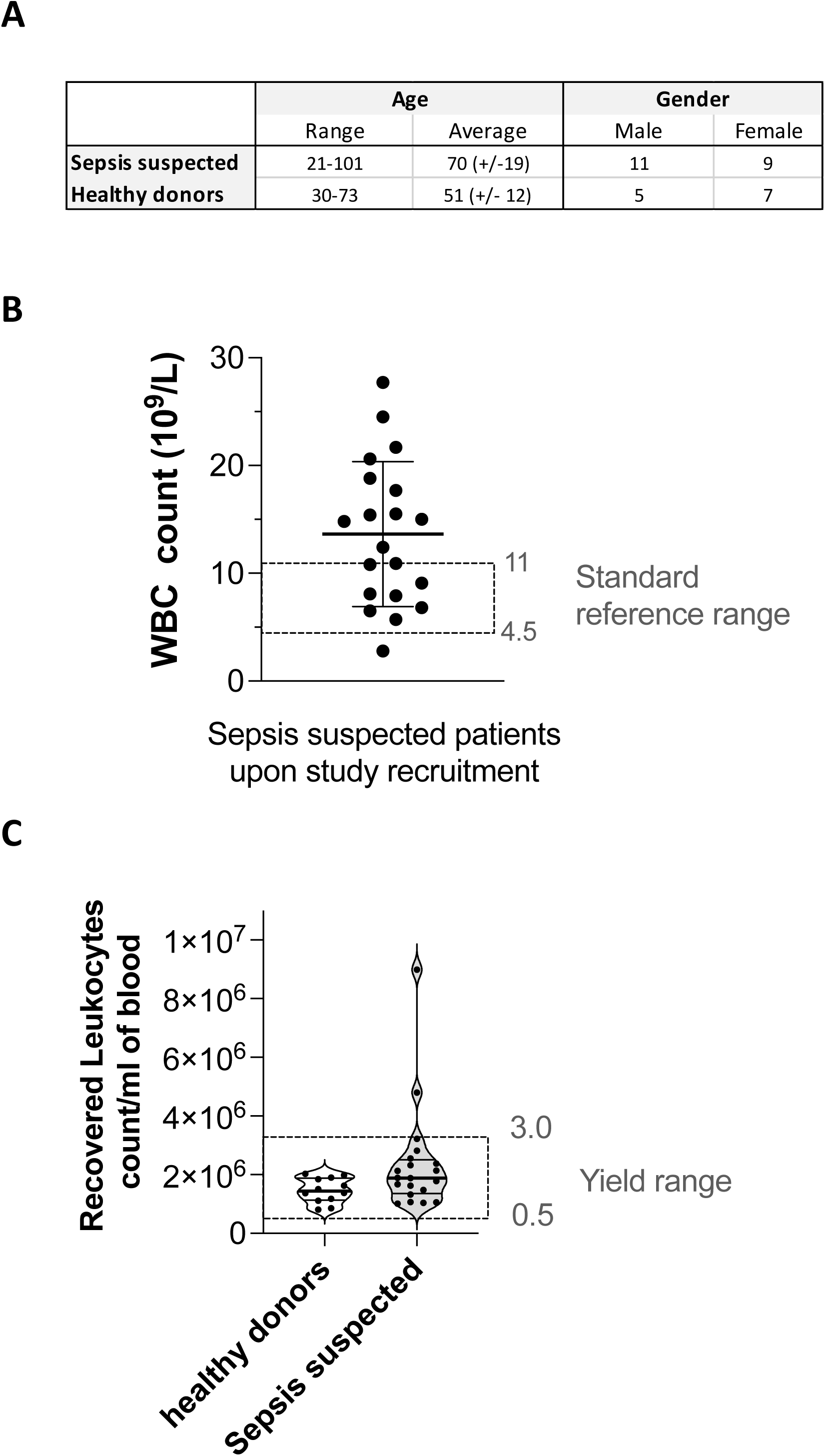
Study cohorts and blood samples used for biological assessment. **A)** Demographics of study participants. **B)** White Blood Cell count for sepsis-suspected study participants measured at time of hospital admission with standard reference range. **C)** Count of leukocytes recovered from the PBMC fraction isolated by density gradient from blood samples of all study participants with the accepted yield range for healthy adults.

### Cytokine and chemokine profiling from blood plasma discriminates healthy controls from sepsis-suspected patients

We performed a flow cytometry-based multiplex analysis, measuring plasma levels for a range of cytokines and chemokines linked to sepsis^21^. Only IL-6, CXCL9, and CXCL10 showed statistical differences between healthy and sepsis-suspected samples (Fig.2A), being elevated in sepsis-suspected patients. IL-5 and CXCL5 showed an inverse trend with reduced plasma levels compared to healthy controls, but this was not significant. We also measured soluble CD14 (sCD14) using an ELISA able to detect full sCD14 as well as its cleaved derivative sCD14 subtype (ST), identifying an increase in circulating levels of sCD14 in sepsis-suspected patients (Fig 2B), in agreement with other studies^32^. Principal components analysis (PCA) of the CC data together with sCD14 indicated some grouping of the data with healthy patients clustering together (Fig. 3A(i)). The biplot shows many loadings with similar magnitudes contributing to the spread of sepsis-suspected patients, including TNF, IFNγ and IL-17A, as well as those with greatest separation measure according to Equation [1] (Fig. 3A(ii)). We repeated the analysis using only IL-6, CXCL9, CXCL10, CXCL5, IL-10, CCL11 and sCD14, the variables with separation measure >0.1 and found that the simpler model was able to achieve a similar level of discrimination (Fig. 3B(i)). The loadings show that differences between sepsis-suspected and healthy controls along PC1 is driven by IL-6, IL-10, CXCL9 and CXCL10 whereas differences along PC2 are due to CXCL5 and CCL11 with sCD14 contributing very little. This unsupervised analysis was complemented by partial least squares regression (PLSR) analysis on the full CC data together with sCD14. A correct classification rate of 93.75% in leave-one-out (L-O-O) classification confirmed the separation between profiles from patients with sepsis-suspected and healthy controls with all sepsis patients correctly classified and just two of the twelve healthy control group incorrectly classified (Fig. 3C(ii)). VIP scores showed IL-6, CXCL9, CXCL10 and CXCL5 to be the most important variables in the model (Fig. 3C(iii)).

**Figure 2:**
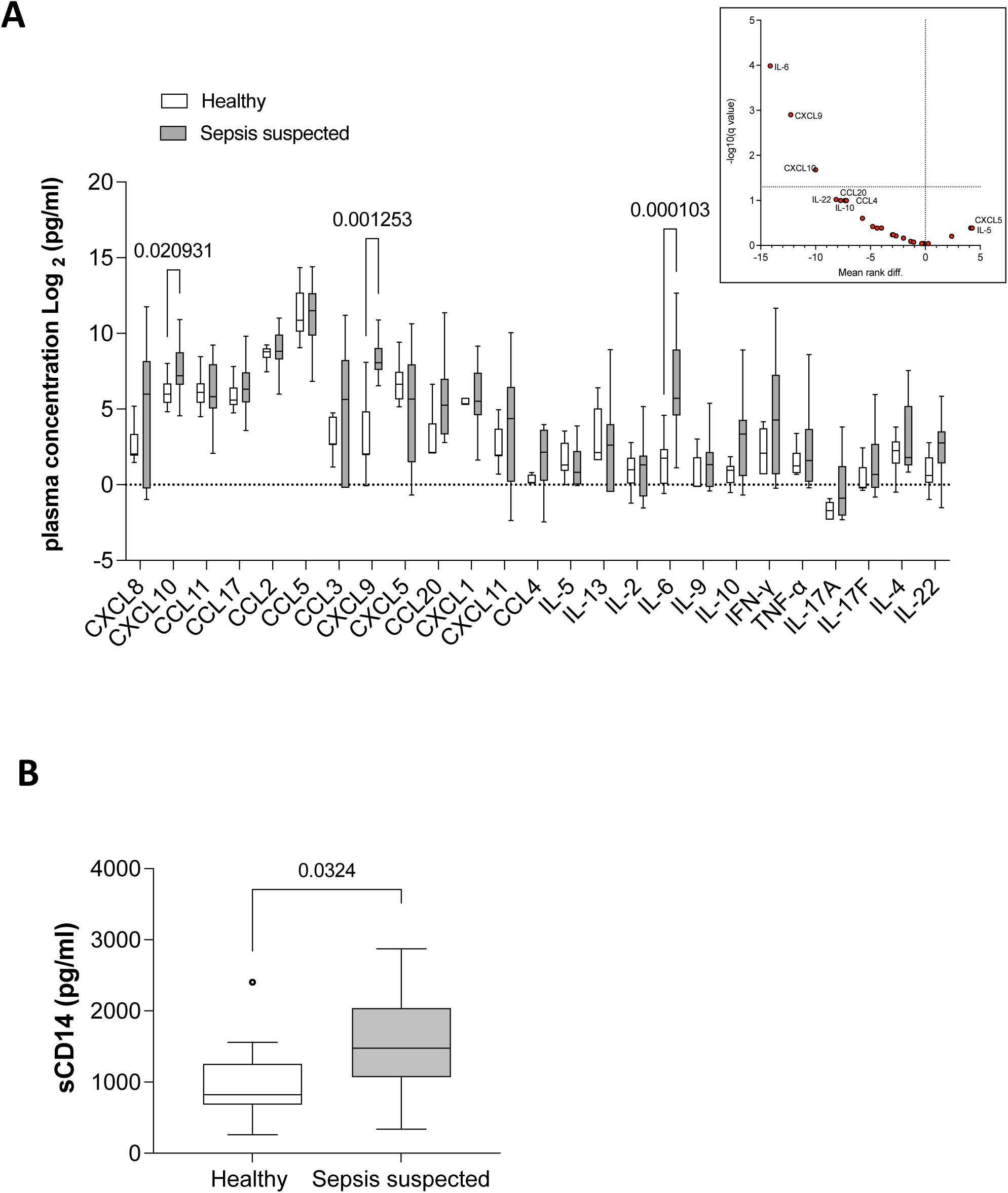
Circulating cytokine and chemokine (CC) signatures for healthy volunteers compared to sepsis-suspected patients. **A)** Box and whisker plot of measured cytokines and chemokines from blood-isolated plasma samples with statistical significance from Mann-Whitney tests using Benjamini, Krieger and Yekutiel adjustment for multiple comparisons. The volcano plot reports the mean rank differences, either increased or decreased, for all markers between the healthy and sepsis-suspected groups. **B)** Box and whisker plot of sCD14 blood concentration measured by ELISA, with statistical significance (p = 0.0324) between healthy and sepsis-suspected groups defined using a Mann-Whitney test.

**Figure 3:**
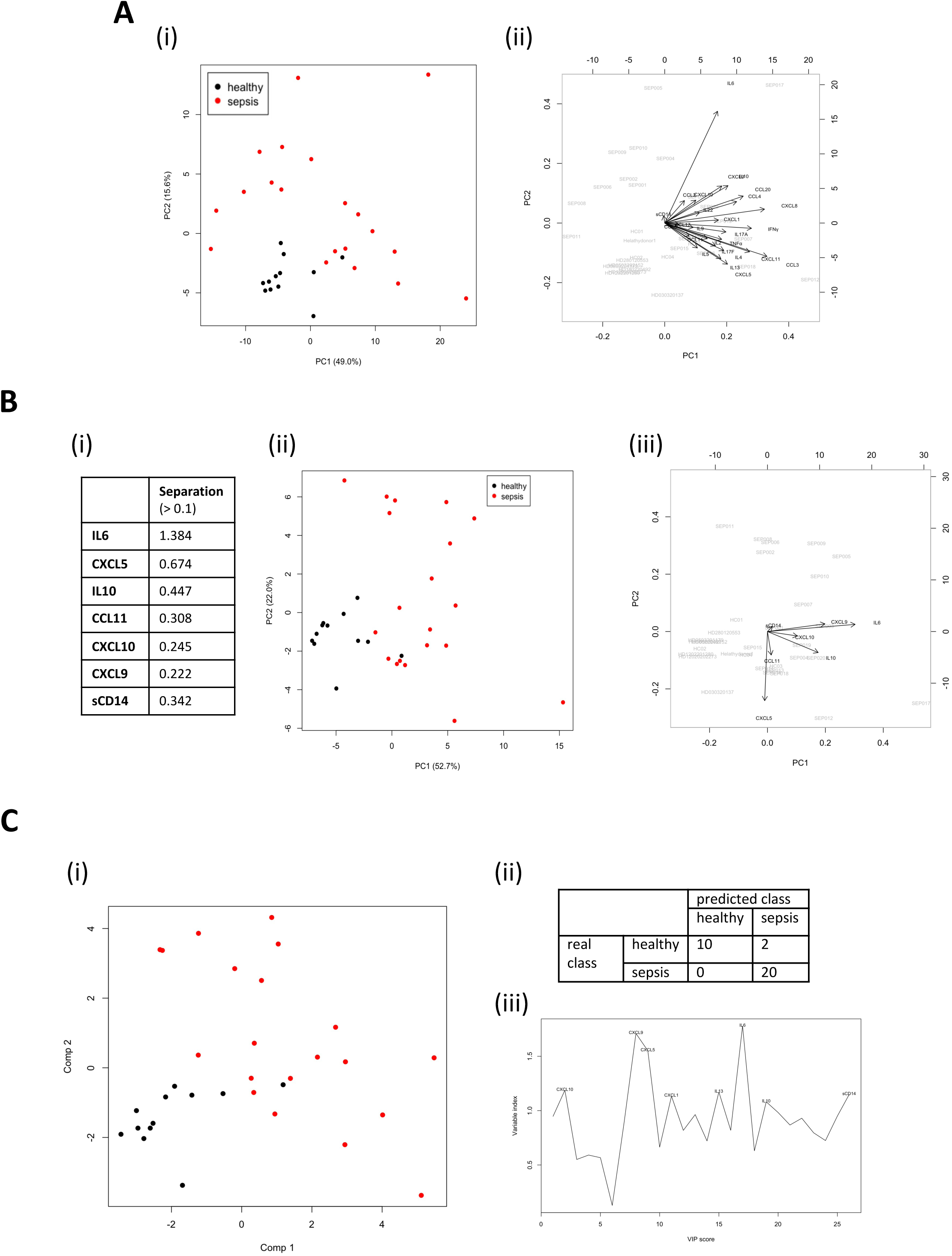
Integration of sCD14 plasma levels with cytokine and chemokine (CC) profiles. **A)** Principal component analysis (PCA) scores plot (i) for the first two components for cytokine and chemokine datasets with sCD14 with biplot (ii) showing the loadings as vectors (black arrows) and scores by sample names in grey. **B)** PCA restricted to variables with separation scores > 0.1 (i), with scores plot (ii) and biplot (iii) showing the loadings. Here, separation is defined by the between groups variance divided by the within groups variance (equation (1), see material and methods). **C)** Partial least squares regression (PLSR) scores plot (i) for the first two latent variables obtained using scaled sCD14 and CC data. The confusion matrix (ii) shows the results obtained using leave-one-out cross validation on scaled data. (iii) The Variable Importance in Prediction (VIP) graph highlighting the most important variables in the data. The dotted line shows the threshold, VIP score = 1, above which the variables are labelled.

Spearman correlation analysis was performed to determine whether specific relationships existed between individual variables that could explain the dominance of IL-6, CXCL9 and CXCL10 in our findings. The heatmaps in Figure 4 show differences when the same variables are considered for the healthy control or the sepsis-suspected group. Distinct clusters of positively correlated variables emerged for the two groups, with no strong correlation uniting IL-6, CXCL9 and 10 within a particular cluster, even for the sepsis-suspected group. Note that few anti-correlations were observed, including IL-5 with many CC and sCD14 in the healthy group, and sCD14 with many CC for sepsis-suspected patients. Overall, the clusters of pairwise correlations do not connect the discriminatory variables separating healthy from sepsis, suggesting that multiple unrelated immune events lead to these blood signatures. The dominant correlation cluster for sepsis-suspected patients, which includes IFNγ, TNF, IL-2, IL-4, IL-5, IL-13, IL-17A/F denotes a mixed Th1, Th2 and Th17 signature.

**Figure 4:**
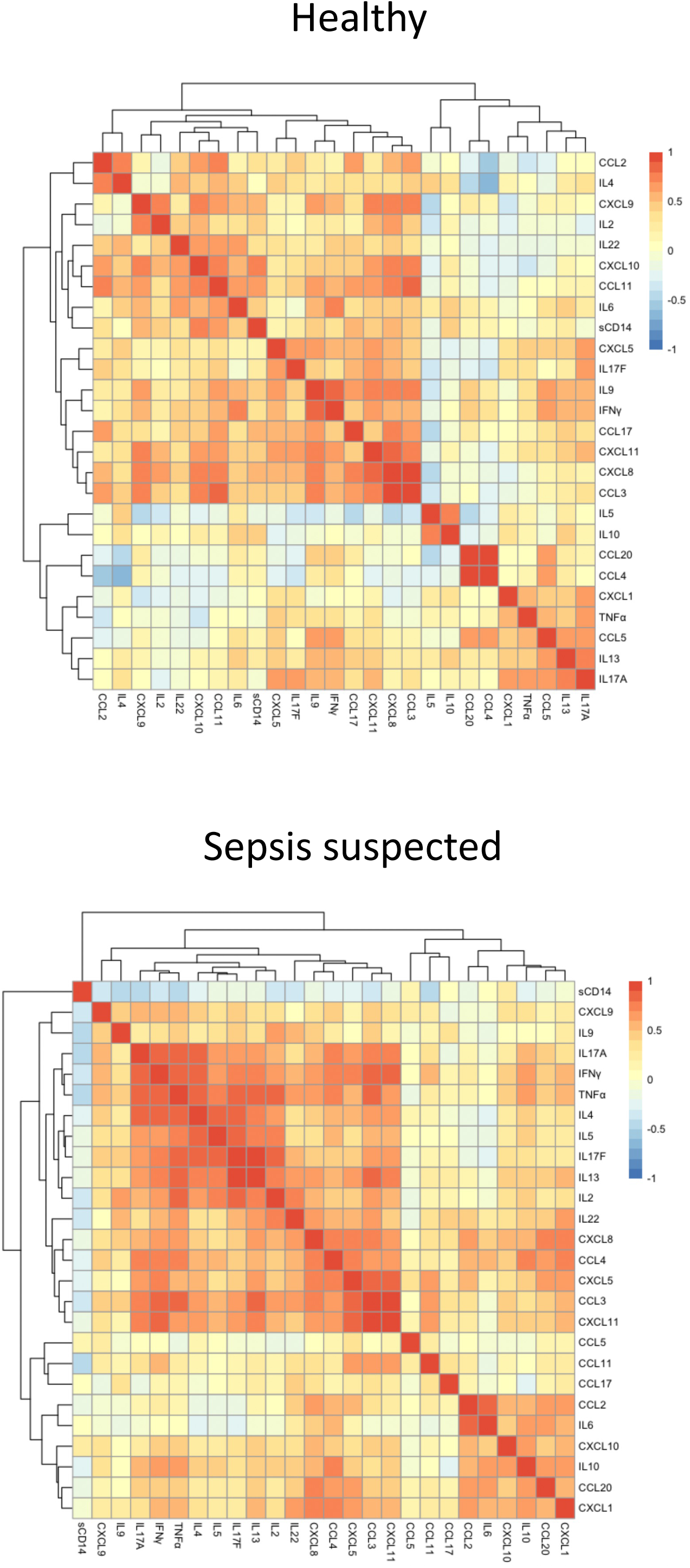
Heatmaps showing Spearman correlation between the different cytokines, chemokines and sCD14, for the healthy control and sepsis-suspected groups.

### Leukocyte changes in sepsis-suspected patients

To compliment the above analysis of chemokines and cytokines, we used a blood cell panel (BCP) of antibodies (Fig. S2) to quantify innate and adaptive immune cell subpopulations in density gradient-isolated blood leukocytes (Fig. 5 & 6), size and granularity for differential gating on granulocytes, monocytes and lymphocytes (Fig. S3A). A further gate was included to capture atypical large monocytes, a population that has been reported expanding in diseases such as COVID-19 and COPD^33, 34^. Analysis across all participant samples showed that leukocytes from sepsis-suspected patients had a significant increase in percentages of low-density granulocytes and regular monocytes coinciding with a reduced frequency of lymphocytes (Fig. 5A), all of which represent early predictors of sepsis^35^. We found no significant differences between the percentage of granulocytes, monocytes and lymphocytes expressing CCR1, CCR2, CCR5, CCR7 or TLR2 (a pattern recognition receptor reported to be specifically affected in sepsis^36^), probably due to the large heterogeneity of expression between individuals (Fig. 5B).

**Figure 5:**
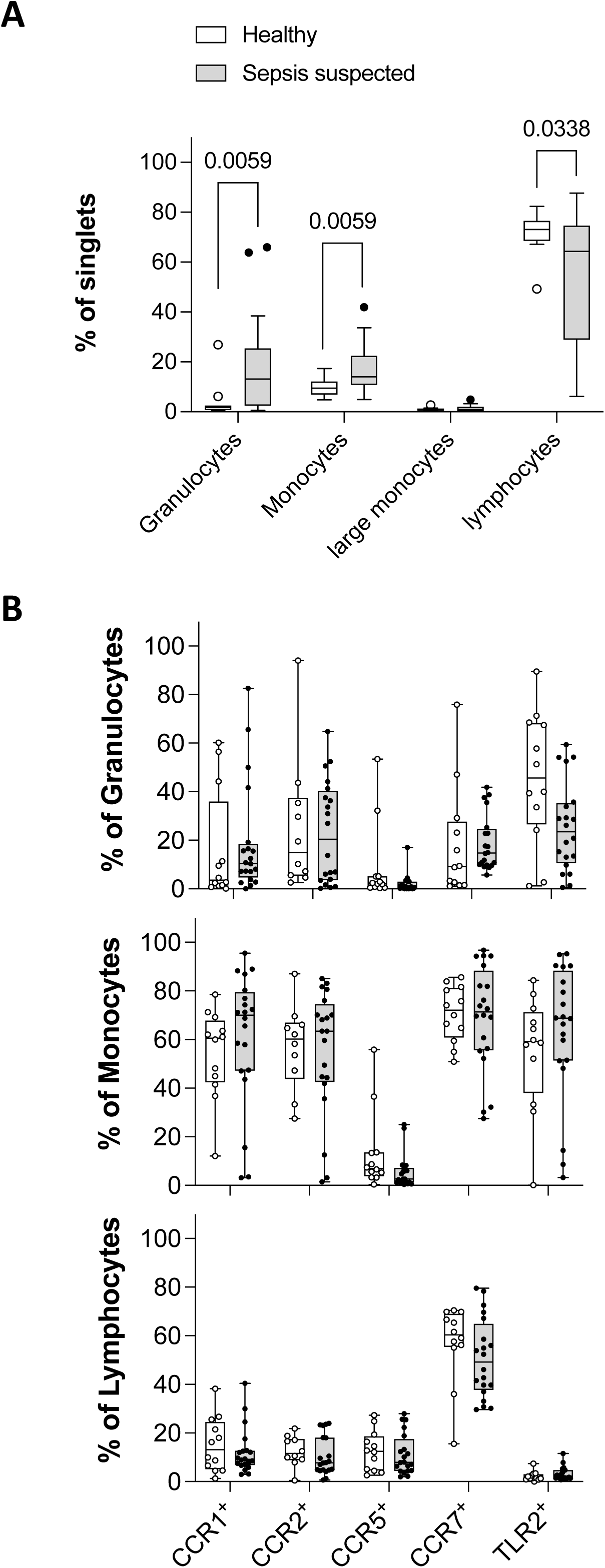
Blood-isolated leukocytes and expression profiles of selected inflammatory cell-surface markers. **A)** Representation of leukocyte sub-populations identified by flow cytometry based on forward and side scatter profiles, comparing healthy volunteers to sepsis-suspected patients. Results are expressed as the percentage of total single cells (singlets) recorded by the cytometer using the mean value from n=5 samples run for each individual. The boxplot shows the interquartile range with outliers using the Tukey method. Statistical significance (adjusted p values) from Mann-Whitney tests for multiple comparisons using Benjamini, Krieger and Yekutiel secondary test. **B)** Expression of CCR1, 2, 5, 7 and TLR2 within the three main leukocytes sub-populations assayed by antibody staining and flow cytometry analysis, defining the percentage of cells positive for the indicated marker. Boxplots for both healthy and sepsis-suspected groups are shown with all points plotted. Mann-Whitney tests showed no statistical differences.

When gating on subsets markers detected by the BCP (Fig. S1), namely the monocytic marker CD14, the T cell receptor CD3 and the B cells marker CD19 (Fig. 6), we confirmed the reduced frequency of lymphocytes, which affected both CD3^+^ T and CD19^+^ B cells in sepsis-suspected patients (Fig. 6A). We found a trend but no significant increase in the percentage of blood CD14^+^ cells, which was not confirmed by restricting our gate to CD14^+^ monocytes (Fig. S3A). Analysing chemokine receptors and TLR2 expression within each of the subset revealed a significant reduction in the proportion of CCR5-expressing CD14^+^ cells and CCR7-expressing in sepsis-suspected patients (Fig. 6B). Interestingly CCR1 and CCR2, which are expressed from genes located in the same cluster as CCR5 on human chromosome 3p21 were not affected^37^. There was no difference in the frequency of CD3^+^ cells expressing these markers apart from a small but significant increase in TLR2 positive cells in sepsis-suspected patients. Finally, sepsis-suspected patients had an increased frequency of CD19^+^ CCR2^+^ cells and a decrease in frequency of CD19^+^ CCR7^+^ cells (Fig. 6B). Of note, combining healthy controls and sepsis-suspected patients, we observed an anti-correlation between the frequency of CCR2- and CCR7-expressing CD19^+^ B cells (Fig. 6C). As monocytes are heterogeneous^38^, we used CD14 and CD16 expression to gate separately on classical, intermediate, and non-classical monocytes (CM, IM and NCM, respectively; Fig. S3B). NCM showed a significant reduction in the frequency of CCR2^+^ cells in sepsis-suspected patients, whereas the frequency of CCR5^+^ monocytes belonging to each subpopulation was reduced in sepsis-suspected patients (Fig. 6D). No differences in the frequency of CCR1, CCR2 and TLR2 positive cells were observed in any monocyte subset (Fig. S4). This flow cytometry analysis indicates that specific but discrete phenotypic changes affecting cells from both the innate and adaptive immune system can separate ED-admitted patients suspected of sepsis from healthy controls. PCA analysis on all parameters measured by multicolour flow analysis with the Blood Cell Panel of antibodies showed some clustering of the healthy controls away from sepsis-suspected patients but the grouping was less clear than seen for the CC data with more spread of the healthy group (Fig. S5). Variable importance in projection (VIP) scores from PLSR model on this data (Fig. 7) showed group separation due to parameters such as CD3, CD14/CCR5, CD19/CCR7 and CD19/CCR2 (Fig. 7B) with a L-O-O classification rate of 87.5%, having two errors for each group (Fig. 7C)

**Figure 6:**
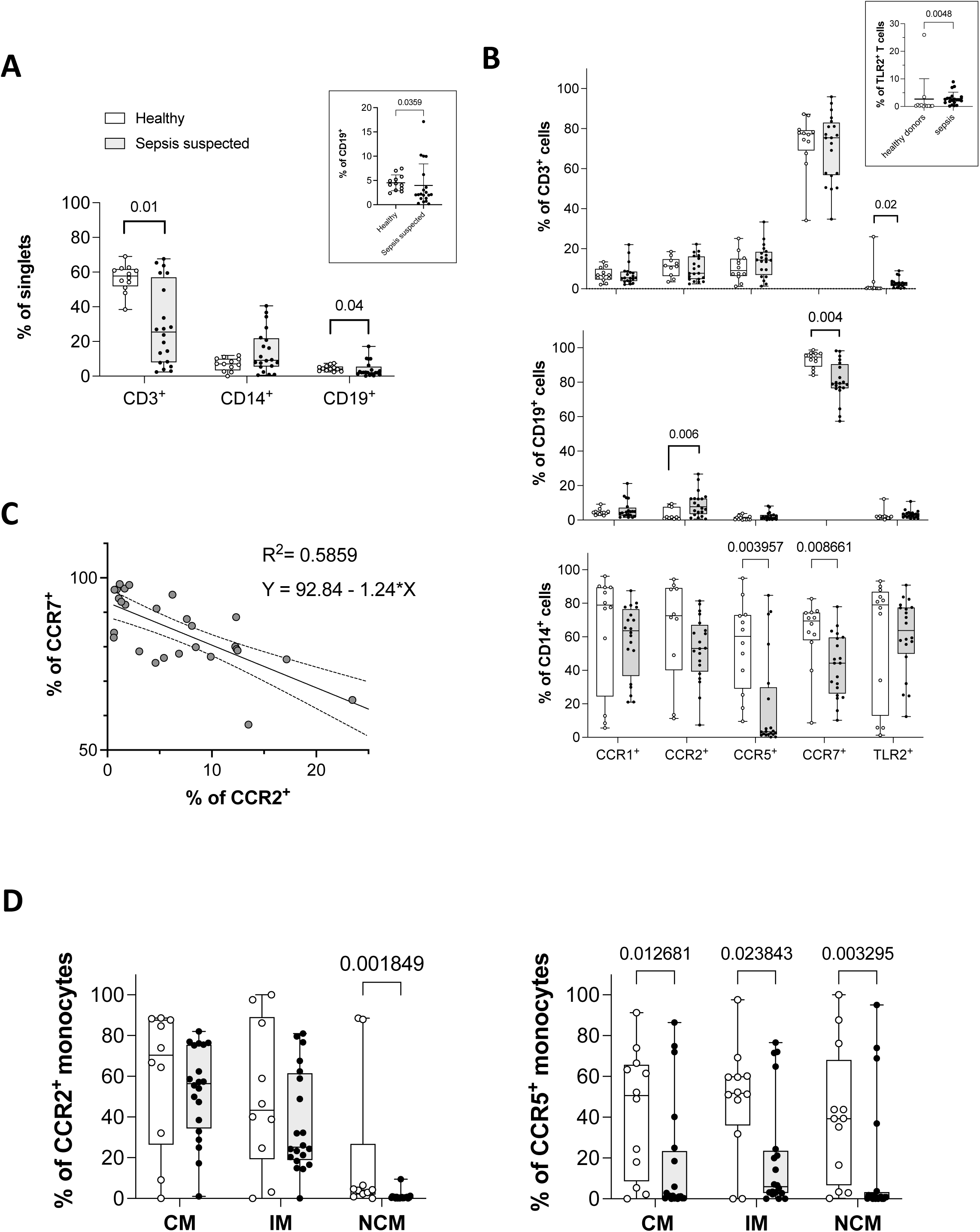
Expression profiles of selected inflammatory cell-surface markers on lymphocyte and monocyte sub-populations. All presented results comparing healthy and sepsis-suspected groups report on the percentage of cells positive for the indicated markers as measured by multicolour flow cytometry analysis using our defined blood cell panel (BCP). **A)** Frequency of T cells, monocytes, and B lymphocytes among single cells recorded by the cytometer (singlets) and based on CD3, CD14 and CD19 expression, respectively; insert shows scatter plot for CD19^+^ distribution with Mann-Whitney test. **B)** Expression of CCR1, 2, 5, 7 and TLR2, on CD3^+^, CD14^+^ and CD19^+^ cells; insert shows scatter plot for TLR2^+^ distribution on CD3^+^ cells with Mann-Whitney test. **C)** Negative correlation between CCR7 and CCR2 expression on CD19^+^ B lymphocytes (Spearman correlation r = -0.6776). The linear regression equation and goodness of fit coefficient (R^2^) are shown, p < 0.00001. **D)** Frequency of classical (CM), intermediate (IM) and non-classical (NCM) monocytes subpopulations, as defined in supplementary figure S4B. For all Boxplot shown with all points, statistical significance was determined using Mann-Whitney tests with adjustment for multiple comparisons (adjusted p values) using Benjamini, Krieger and Yekutiel secondary tests.

**Figure 7:**
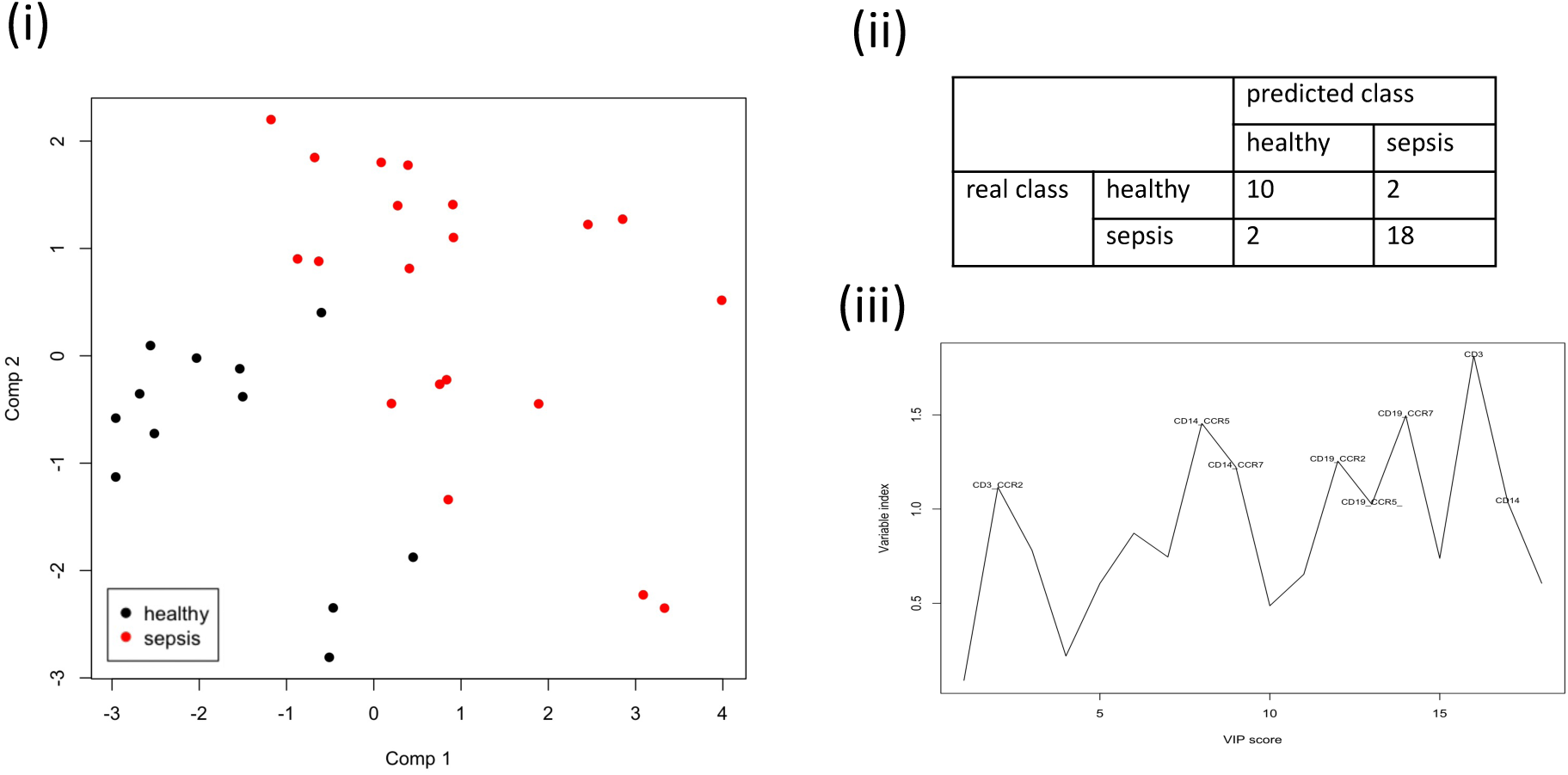
Separation of healthy and sepsis-suspected profiles based on the different blood cell panel (BCP) parameters analysed. The partial least squares regression scores plot for the first two latent variables (i) obtained using the subset of BCP variables shows clear separation between the healthy and sepsis-suspected groups. Accuracies showed in the table were obtained using a leave-one-out cross validation on scaled data (ii) and the VIP scores indicate the importance of predictor variables (iii; with VIP scores >1.0 labelled).

### T cell activation profiles in sepsis-suspected patients

T cells are essential mediators of the host response to sepsis, but recent studies have also indicated that T cell dysregulation impaired this response^39, 40, 41^. This includes all main T cells subtypes from effector CD4 and CD8, regulatory (Treg) and memory T cells^42, 43, 44^. To assess whether significant T cells changes can be detected in blood samples from our sepsis-suspected patients, we performed multiplex flow cytometry analysis using a T cell activation panel (TCAP; Fig. S1). Gating to identify CD4^+^ and CD8^+^ cells within the lymphocytes gate, we first noticed that CD8 spread between CD8^+^(high) and CD8^+^(low) cells (Fig. 8A). CD8^+^(high) correspond to classical CD8^+^ T cells, while CD8^+^(low) have been reported as a distinct subpopulation of activated CD8 effector cells in human peripheral blood^45^. Comparing the representation of CD4^+^ and CD8^+^ populations in lymphocytes of healthy controls and sepsis-suspected patients, we showed a highly significant reduction in CD4^+^ T cells associated with sepsis (Fig. 8B). From the percentage of CD4^+^ and CD8^+(high^ ^&^ ^low)^ lymphocytes we calculated the CD4/CD8 ratio, which for healthy individuals is expected to be greater than one but reduces with age^46^. The average value was significantly lower for the sepsis-suspected group, with many patients presenting an abnormal inverted (<1) CD4/CD8 ratio (Fig. 8C).

**Figure 8:**
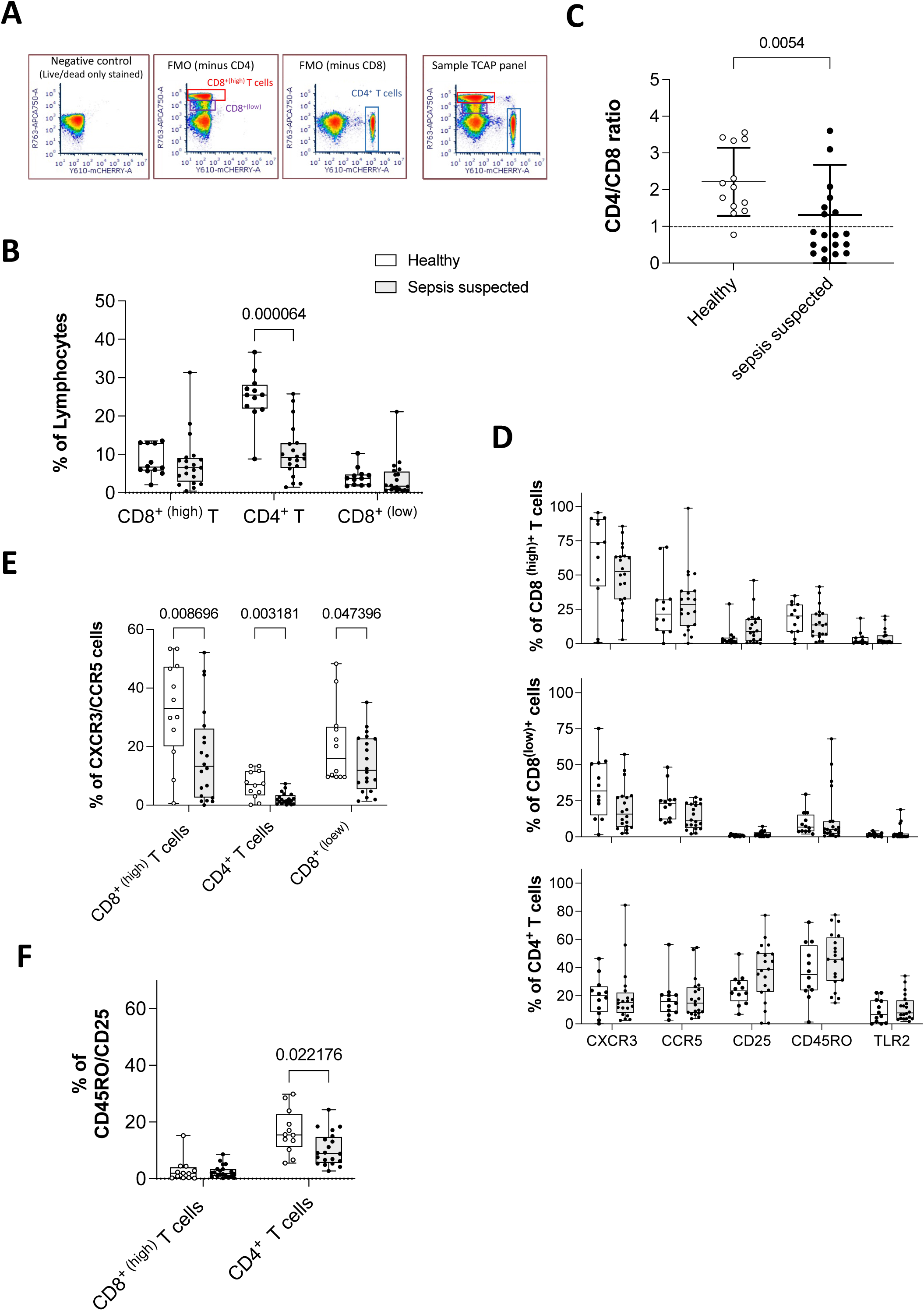
Expression profiles of activation markers on T cells subpopulations. All Boxplot with all points comparing healthy and sepsis-suspected groups, report on the percentage of cells positive for the indicated markers as measured by multicolour flow cytometry analysis using our defined T cell activation panel (TCAP). **A)** Flow strategy using fluorescence minus one (FMO) samples to gate CD4 and CD8 lymphocytes [CD4^+^ T, CD8^+^ (high) T and CD8^+^ (low) cells]. **B)** Frequency of CD4 and CD8 cells among gated lymphocytes. **C)** Scatter plot comparing CD4/CD8 cell ratios in each group (dotted line marks normal ratio >1.0). **D)** CXCR3, CCR5, CD25, CD45RO and TLR2 expression on CD4^+^ T, CD8^+ (high)^ T and CD8^+(low)^ cells. **E)** CXCR3 and CCR5 co-expression on CD4^+^ T, CD8^+ (high)^ T and CD8^+(low)^ cells. **F)** CD45RO and CD25 co-expression on CD4^+^ and CD8^+ (high)^ T cells. Statistical significances were determined using Mann-Whitney tests with adjustment for multiple comparisons (adjusted P-values) by Benjamini, Krieger and Yekutiel secondary test.

For each type of T cell, we assessed the percentage of cells co-expressing two chemokine receptors CXCR3 and CCR5 as markers of T cell activation^47, 48^, CD25 as a marker of activated T cells and Tregs^49^, the CD45RO T cell memory marker^50^, and TLR2 as a regulator of T cell activation in response to infection^51, 52^. Based on the gating strategy described in Fig. S6. No significant difference was seen when assessing the expression of each marker individually (Fig. 8D). However, when assessing combinations of markers, we found a loss of CXCR3/CCR5 dual expression on CD4^+^ and CD8^+^ T cells in sepsis-suspected patients (Fig. 8E), a phenotype reported for Th1-associated T effector/memory (TEM) cells linked with inflammatory reactions^53, 54, 55, 56^. The specific loss of CXCR3/CCR5 double positive T cells was independent from which other markers were expressed (Fig. S7A & B)

In addition, co-expression analysis of CD45RO and CD25 revealed a significant reduction with sepsis for CD4^+^ T cells but not their CD8^+^ counterparts (Fig. 8F). These CD4^+^ CD45RO/CD25 double positive cells have been reported as Type-1 like regulatory T cells^57, 58, 59^. Finally, TLR2 expressing T memory cells (CD45RO^+^)^60^ were comparably represented in samples from healthy controls and patients (Fig. S7D).

PCA analysis using the parameters measured with the TCAP did not separate healthy controls and sepsis-suspected groups (Fig. S8). Overall, the TCAP analysis showed that lymphopenia is driven by a loss of CD4^+^ T cells and that remaining T cells are deficient in the T memory and regulatory cell compartments detectable through CXCR3/CCR5 and CD45RO/CD25 co-expression.

### Analysis of integrated parameters from all datasets

Having considered individual datasets, we also analysed the variables from all datasets together (Table S2). To determine the parameters that dominate the changes in our sepsis-suspected cohort, we used two supervised methods, PLSR (Fig. 9A) and Random Forest (Fig. 9B), that allow the most important variables to be identified. Both methods achieved good discrimination between healthy controls and sepsis-suspected patients with correct classification rates of 93.8% and 90.6% respectively for PLSR (Fig. 9A (ii)) and Random Forest (Fig. 9B (i)). The most discriminatory variables were identified from VIP scores in PLSR (VIP>1.3, Fig. 9A (iii)) and according to their importance in Random Forest classification based on mean decrease in accuracy (Fig. 9B (ii)). Although there are differences between the two methods, some variables, such as IL-6, CXCL9, CXCL10, CCR5^+^ monocytes, CD4^+^ and CXCR3/CCR5 co-expressing T cells emerge as important parameters in both analyses.

**Figure 9:**
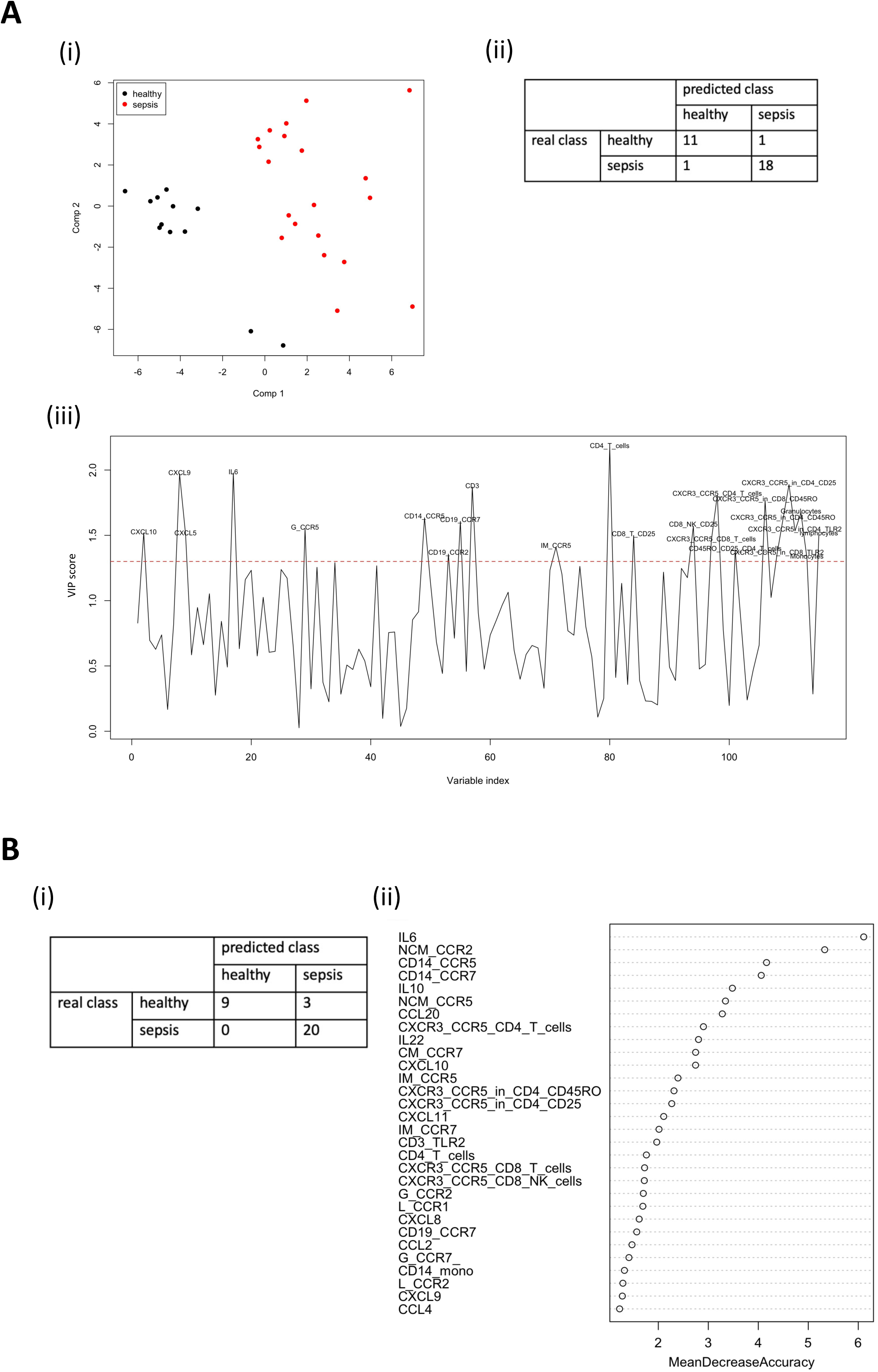
Separation of healthy and sepsis-suspected profiles based on variables measured across all panels. **A)** Partial least squares regression (PLSR) results from model obtained with all variables. (i) The PLS scores plot generated from all data shows clear separation between healthy (red) and sepsis-suspected (black) profiles. (ii) The results from L-O-O cross-validation show one classification error for each group. (iii) VIP scores graph highlighting the most important predictor variables in the PLS model. The dotted line shows the threshold, VIP score = 1.3, above which the variables are labelled. **B)** (i) Confusion matrix showing the results from Random Forest classification using L-O-O cross-validation. (ii) The most discriminatory variables sorted by importance based on mean decrease in accuracy.

### Blood immune signatures and clinically validated sepsis status of patients

In order to assess whether this could come from heterogeneity in the sepsis-suspected group, we revisited our analysis in view of the clinical diagnoses at discharge for all but one patient who withdrew in the later phase of our study. This created two subgroups of patients whether they were assessed as confirmed (N=13) or unconfirmed (N=6) cases of sepsis (see Table S1).

The possible correlation between parameters identified as discriminatory in PLS analysis was investigated within individual groups and subgroups from our study to determine whether correlations were maintained or disturbed by sepsis (Fig. 10). For healthy controls the heatmap shows a contrasting pattern of positive and negative correlations, with for example a cluster including inflammatory soluble (IL-6, CXCL9, CXCL10 and sCD14) and cellular parameters (CCR5 expression on monocytes or granulocytes, CCR2 on CD19^+^ B cells) being anti-corelated with CXCR3/CCR5 T cells. This pattern is lost in the sepsis-suspected group with very few parameters showing any relationship. The separate analysis of unconfirmed and confirmed sepsis cases led to the reappearance of a contrasting pattern for the unconfirmed group, although different from healthy controls. For example, IL-6 and CCR5 expression on CD14^+^ monocytes appear anti-correlated with CXCL9, while sCD14 opposes CXCR3/CCR5 T cells. As with the sepsis-suspected group, the heatmap for confirmed sepsis shows less of a pattern, but some clustering of CXCR3/CCR5 T cells and with IL-6, CXCL10 and IL-10 positively correlated. Interestingly, both healthy controls and unconfirmed sepsis groups showed negative correlation between sCD14 and CXCR3/CCR5 T cells, which is not the case for the sepsis-confirmed group. These results suggest that immune blood profiling can expose patterns distinguishing early sepsis from other forms of systemic inflammation, with the combination of IL-6, CXCL9, CXCL10 and sCD14 plus CCR5 and CXCR3 expression emerging as a potential biomarker signature.

**Figure 10:**
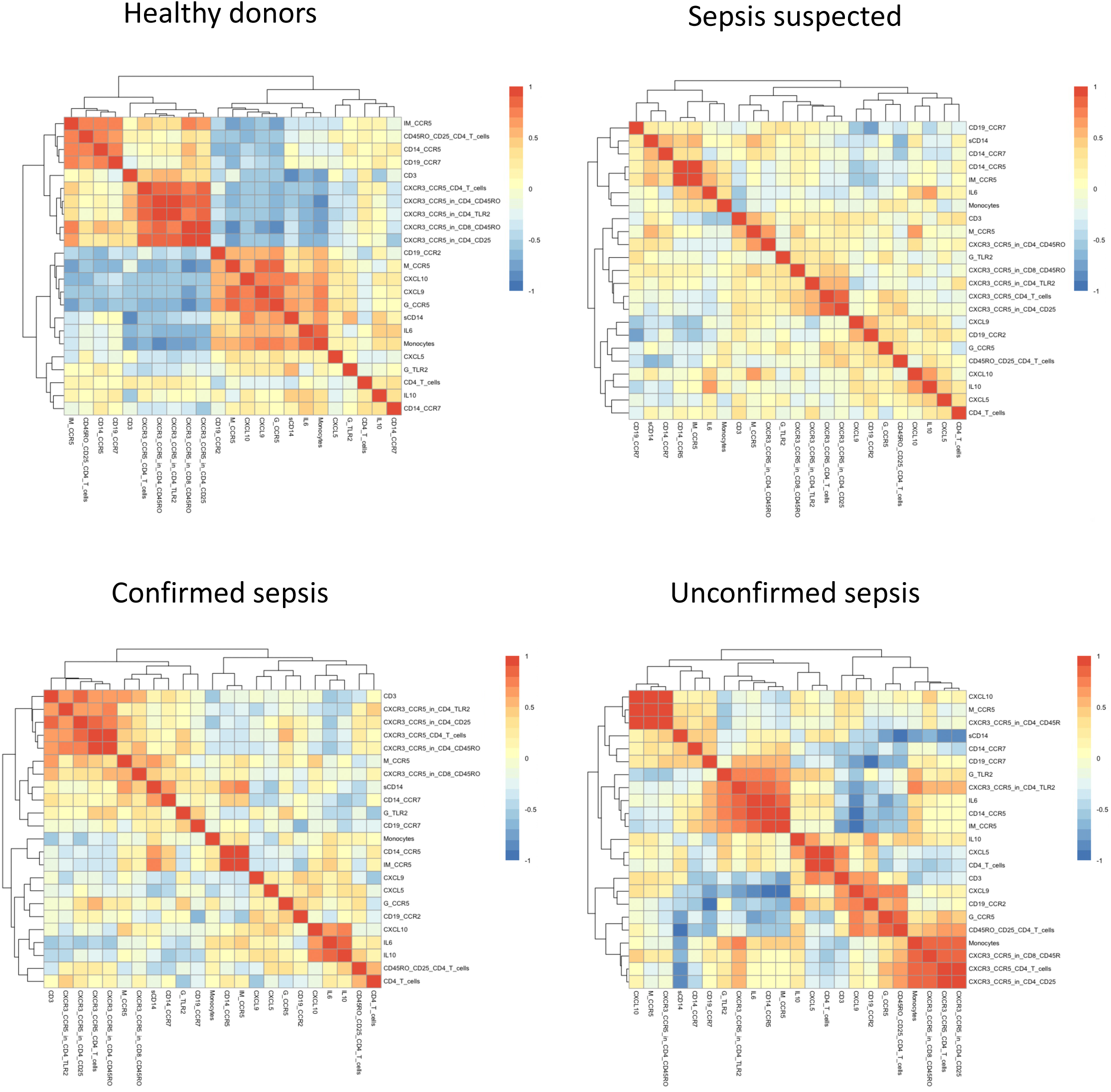
Correlation analysis restricted to the prominent variables identified across the entire dataset. Spearman correlations using the most important predictor variables (VIP >1.3) from PLSR analysis of all measured variables as reported in Figure 9. Heatmaps display corelations within the Healthy, sepsis suspected, confirmed and unconfirmed sepsis subgroups, as indicated.

ANOVA tests on individual parameters from the three different datasets showed some significant differences between the confirmed and unconfirmed subgroups (Fig. 11). For CC data, we found a similar increase in IL-6 and CXCL9 for both subgroups in comparison to the healthy group, but CXCL10 accumulation was only significant for the unconfirmed sepsis group and IL-5 levels are significantly different between the confirmed and unconfirmed subgroups (Fig. 11A). For the BCP dataset, only confirmed sepsis patients exhibited a loss of CCR7^+^ lymphocytes (Fig. 11B) whereas in the TCAP dataset, the CD4^+^ T cells-driven lymphocytopenia was specific for the sepsis confirmed subgroup (Fig. 11C). There was also a significant increase in activated CD8 T cells (CD25^+^) for unconfirmed cases versus confirmed sepsis and healthy controls (Fig. 11D). Interestingly, the severe reduction in CXCR3/CCR5 co-expressing CD4^+^ and CD8^+^(high) T cells specifically affected patients with confirmed sepsis diagnosis (Fig.11E).

**Figure 11:**
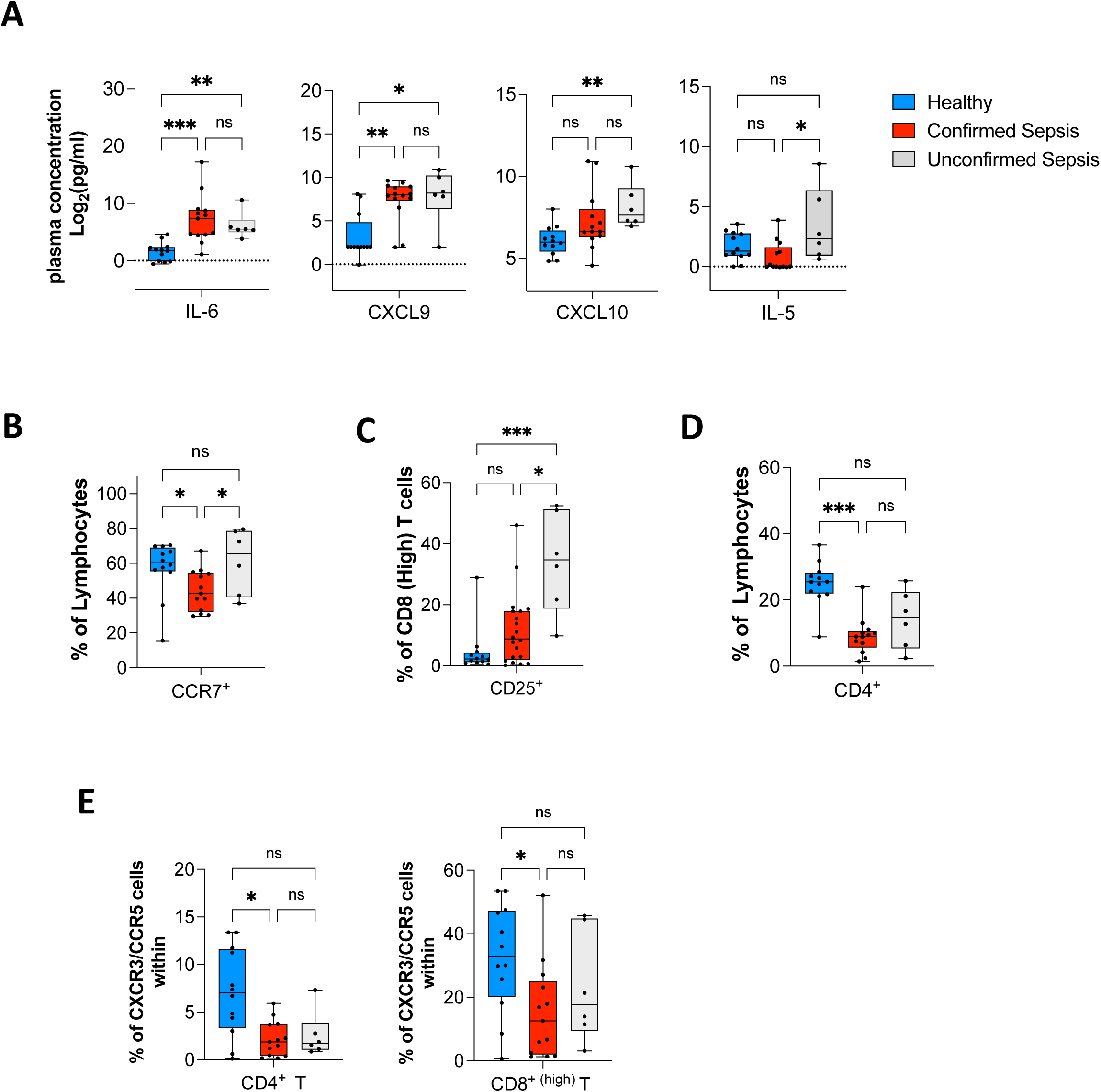
Subgroup analysis by individual markers. Analysis of each marker individually identified a series of variables that show significant differences between the various subgroups, coloured according to healthy individuals (blue), confirmed (red) and unconfirmed sepsis (grey) cases. **A)** Cytokines and chemokines showing significant differences between at least one pair of subgroups. **B)** The percentage of CCR7^+^ lymphocytes, **C)** CD4^+^ lymphocytes, **D)** CD25^+^ CD8^+ (high)^ T cells and **E)** CXCR3/CCR5 double positive cells in CD4^+^ and CD8^+ (high)^ subpopulations. Statistical significance was determined using ANOVA and Kruskal-Wallis multiple comparisons test (adjusted p values: *** p < 0.001, ** p < 0.01, * < 0.05, ns= non-significant).

However, PCA analysis of the full CC and sCD14 dataset did not show separation of confirmed and unconfirmed sepsis patients, as was also the case for the BCP and TCAP datasets (Fig. S9A). Separation was also not possible when combining all three datasets, despite patients segregating away from healthy controls (Fig. S9B). Our analysis suggests that blood signatures based on analysis of many parameters covering innate and adaptive immunity could not discriminate sepsis from other conditions with systemic inflammatory responses.

## Discussion

Prompt recognition and accuracy in detecting sepsis in ED is crucial to rapidly administer antibacterial agents and treatments that can reduce mortality. However, in the initial stage sepsis presentation often resembles non-infectious SIRS associated with a range of conditions such as trauma, ischaemia or autoimmune disorders^61^. In this pilot observational study we combined peripheral blood profiling for circulating inflammatory markers, leukocytes phenotyping and data integration using mathematical methods to expose signatures of altered immune response that could distinguish sepsis cases upon hospital admission. Overall, we show that integrated profiles of dominant measured variables contrasted between healthy controls, patients admitted for sepsis-suspected, and patients with a later confirmed or unconfirmed clinical diagnosis of sepsis (Fig. 10).

We found that blood circulating biomarkers could segregate patients from healthy controls (Fig. 2 & 3), with significant accumulation of IL-6, CXCL9 and CXCL10 in sepsis-suspected patients (Fig. 2A) supported by qualitative changes in sCD14, IL-10, CCL11 and CXCL5 (Fig. 3). IL-6 is produced by macrophages as well as T cells and a known mediator of the acute phase of responses to infection, but do not differentiate sepsis from non-infectious SIRS^62^. CXCL9 and CXCL10 represent novel markers with CXCL10 being more dominant in the unconfirmed group (Fig. 11A). Although, these IFNγ−induced chemokines that interact with CXCR3 to elicit immune responses and recruit immune cells to inflamed/infected organs^63^, were also upregulated in non-bacterial systemic infection like COVID-19^64^. Interestingly, the overall CC blood profiles and pairwise correlation patterns for patients with suspected, confirmed or unconfirmed sepsis (Fig. 2A, 4 and S9) are distinct from what we previously reported for COVID-19 patients admitted to ED, and early sepsis was not marked by a cytokine storm^64, 65, 66^. However, the dominant cluster of positive correlations included IFNγ, TNFα, IL-2, IL-4, IL-5, IL-13 and IL-17A/F (Fig. 4 and S9) indicating changes in Th1, Th2 and Th17 profiles and reflecting early immune alterations thought to be linked with sepsis severity^67, 68, 69^

Leukocytes immunophenotyping complemented the soluble inflammatory marker screen, looking for cellular changes that could be correlated with biomarkers profiles and reflect specific effects associated with early sepsis-induced changes and imbalance of the immune network^70^. Unlike many published studies, we immunophenotyped freshly isolated and unfixed leukocytes since the process of fixation is known to impede the detection of cell surface markers particularly chemokine receptors^71, 72, 73^. Generally, patients suspected of sepsis presented with high white blood cell counts and significant changes for composition due to an increase in number of granulocytes and monocytes that coincided with a loss of lymphocytes (Fig. 5A). The apparent lymphopenia was driven by T and B cells (Fig. 6A), with a prominent loss of CD4^+^ T cells resulting in abnormal CD4/CD8 ratios (Fig. 8C). A decline in CD4^+^ T cells is well-documented for sepsis^74^ and loss of CD4 lymphocytes could separate confirmed from unconfirmed cases in our study (Fig. 11D). Interestingly B cells also showed altered expression of the chemokine receptors CCR2 and CCR7 in the sepsis suspected group (Fig. 6B). For all individuals there was an inverse relationship between the level of CCR2 and CCR7 at the surface of blood B cells (Fig. 6C). This observation may be explained by CCR2 being present on immature B cells and downregulated with maturation, while CCR7 is a known marker of naïve and mature B cells^75, 76^. Therefore, a switch in expression of the two receptors in sepsis-suspected patients could reflect an early alteration in the B cell compartment. Other remarkable changes included the collapse of CCR5 positive monocytes (Fig. 6B & D) and of CXCR3/CCR5 co-expressing CD4^+^ and CD8^+^ T cells (Fig. 8E & 11E, S7). CCR5 positive monocytes have recently been reported to be crucial to control sepsis in a murine model^77^ but their loss did not discriminate confirmed from unconfirmed sepsis cases (data not shown) and may therefore be unrelated to the cause of infection. Conversely, the loss of CXCR3/CCR5 double positive T cells was specifically associated with confirmed cases of sepsis (Fig.11E). There is evidence for CXCR3/CCR5 T cells being associated with infiltration of inflammatory sites and inflammatory reactions^53, 55^, and the loss of these chemokine receptors has been reported to impair T cell response to infection^78^. Interestingly, in our study receptors loss coincided with accumulation of the CXCR3 ligands CXCL9/10 and not CXCL11, which interacts differently with CXCR3 and exert a distinct biological activity^79, 80^. CXCR3 activity has been reported to impact on the development and functioning of both CD4^+^ and CD8^+^ T cell compartments^80, 81, 82^. Mechanistically, it has been shown that ligand-mediated activation of CXCR3 leads to receptor degradation and therefore loss of surface expression, with CXCL9/10 acting preferentially on activated T cells^83^. CXCR3 activation also triggers CCR5 cross-phosphorylation on CXCR3/CCR5 T cells blocking their migration^84^. Therefore, there is a possible direct relationship between CXCL9/10 accumulation and loss of CXCR3/CCR5 T cells in early sepsis. Note that some CD4^+^ CD45RO/CD25 Tregs can also co-express CXCR3 and CCR5^85^ but these rare cells were not affected in our study (Fig. S7C), indicating that the loss of the chemokine receptors is restricted to the Th1-associated TEM T cells. This could be part of a wider loss in Th-1 cell populations, which has been documented for ICU-admitted community acquired sepsis patients^67^.

Overall, our study indicates that even at its earliest stage of detection sepsis causes changes across the innate, adaptive, memory and regulatory compartments compared to healthy controls. Globally, these changes could not separate sepsis from other cases of systemic inflammation (sepsis confirmed vs unconfirmed). The multi-layered aspect of the systemic response meant that by investigating an extensive range of parameters we could separate confirmed cases of sepsis based on correlation patterns of dominant immune variables, but not pin down a unique sepsis biomarker-based signature.

However, this is a pilot observational single-centre study, which as such presents several limitations. The small sample size may bias the results, while patient heterogeneity may muddle some observations. While patients were recruited on the day of their admission to hospital, the precise time of potential sepsis onset could not be ascertained, and the dynamic nature of sepsis progression may mask stage-specific alterations. In addition, only a minority of patients enrolled in our study had a bacterial infection directly confirmed by the hospital microbiology lab tests (Table S1). Finally, sepsis is a condition that prevalently affect elderly individuals and age-related changes in immunity as well as dysfunctions due to existing underlying conditions potentially increase the difficulty in discriminating between early sepsis and other forms of systemic inflammation, even based on extensive blood immune profiling. Note that similar issues were raised in a very recent study using machine learning models to predict mortality among a cohort of 77 sepsis patients, and which identified the frequency of T cells and the expression of CXCR3 on CD4 T cells as dominant parameters^86^.

Nevertheless, our investigations exposed a new axis of dysregulation for CCR5 and CXCR3 in early sepsis. Interestingly, blockade of CXCR3 or expression of CCR5 are protective in experimental animal models of sepsis, and both receptors are required for protective T-cell mediated response to bacteria in mice^77, 78, 87^. The notion that CXCR3 blockade or CCR5 expression had a similar outcome is compatible with the process of CXCL9/10-mediated activation down-modulating CXCR3 also removing CCR5^84^. With these two chemokine receptors driving efficient Th1-type adaptive immunity by influencing the positioning and balance in Tregs, T effector and memory cells during inflammation^88^, the question remains whether the collapse in blood T cells CXCR3/CCR5 we observed is part of the cause or a consequence of sepsis.

## Data Availability

All data produced in the present study are available upon reasonable request to the authors

## ACKNOWLEDGEMENTS

The study was sponsored by York & Scarborough Teaching Hospitals NHS Foundation Trust. It was supported by research priming funds from the University of York and by the Elsie May Sykes Research Award administrated by Research & Development at The York Hospital, which also provided advice and secured local and national approvals. The views expressed in this publication are those of the authors and not necessarily those of the NHS, the National Institute for Health Research, or the Department of Health. We thank staff from York & Scarborough Teaching Hospitals NHS Foundation Trust for sample processing, and staff at the Imaging and Cytometry Lab in the University of York Bioscience Technology Facility for technical support and advice. We thank the Blood and Biofluids service managed by the York Tissue Bank and its manager at the time Dr James Fox for blood samples collection from healthy university volunteers. We are grateful for colleagues from HYMS Experimental Medicine and Biomedicine group for their support, in particular Profs Paul Kaye and Dimitris Lagos for constructive discussions and critically reading the manuscript. Finally, we thank all participants, patients as well as healthy volunteers from York Hospital and the University of York’s cohort who donated blood samples for this study.

## AUTHOR CONTRIBUTIONS

NS, NT and DY designed the study. TJ, NT and DY supervised the clinical side of the study including collection and analysis of clinical data; RC oversaw the recruitment with informed consent of patients and healthy volunteers from the hospital and blood sample collection; GF was the study clinical manager. DK and NS performed the biological experiments, KH contributed to the flow cytometry analyses, and JW performed the statistical analyses. DK, JW and NS were responsible for data interpretation and the writing of the manuscript.

## References

1. Rudd, K.E. et al. Global, regional, and national sepsis incidence and mortality, 1990-2017: analysis for the Global Burden of Disease Study. Lancet 2020:395, 200–211. 10.1016/S0140-6736(19)32989-7.

2. Husabo, G. et al. Early diagnosis of sepsis in emergency departments, time to treatment, and association with mortality: An observational study. PLoS One 2020:15, e0227652. 10.1371/journal.pone.0227652.

3. Liu, V.X. et al. The Presentation, Pace, and Profile of Infection and Sepsis Patients Hospitalized Through the Emergency Department: An Exploratory Analysis. Crit Care Explor 2021:3, e0344. 10.1097/CCE.0000000000000344.

4. Vincent, J.L. The Clinical Challenge of Sepsis Identification and Monitoring. PLoS Med 2016:13, e1002022. 10.1371/journal.pmed.1002022.

5. Litell, J.M., Guirgis, F., Driver, B., Jones, A.E. & Puskarich, M.A. Most emergency department patients meeting sepsis criteria are not diagnosed with sepsis at discharge. Acad Emerg Med 2021:28, 745–752. 10.1111/acem.14265.

6. De Backer, D. & Dorman, T. Surviving Sepsis Guidelines: A Continuous Move Toward Better Care of Patients With Sepsis. JAMA 2017:317, 807–808. 10.1001/jama.2017.0059.

7. Opota, O., Croxatto, A., Prod’hom, G. & Greub, G. Blood culture-based diagnosis of bacteraemia: state of the art. Clin Microbiol Infect 2015:21, 313–322. 10.1016/j.cmi.2015.01.003.

8. Punyadeera, C. et al. A biomarker panel to discriminate between systemic inflammatory response syndrome and sepsis and sepsis severity. J Emerg Trauma Shock 2010:3, 26–35. 10.4103/0974-2700.58666.

9. Barichello, T., Generoso, J.S., Singer, M. & Dal-Pizzol, F. Biomarkers for sepsis: more than just fever and leukocytosis-a narrative review. Crit Care 2022:26, 14. 10.1186/s13054-021-03862-5.

10. Tang, J. et al. Serum IL-6 and procalcitonin are two promising novel biomarkers for evaluating the severity of COVID-19 patients. Medicine (Baltimore) 2021:100, e26131. 10.1097/MD.0000000000026131.

11. Kataja, A. et al. Kinetics of procalcitonin, C-reactive protein and interleukin-6 in cardiogenic shock - Insights from the CardShock study. Int J Cardiol 2021:322, 191–196. 10.1016/j.ijcard.2020.08.069.

12. Mosevoll, K.A. et al. Inflammatory Mediator Profiles Differ in Sepsis Patients With and Without Bacteremia. Front Immunol 2018:9, 691. 10.3389/fimmu.2018.00691.

13. Frimpong, A. et al. Cytokines as Potential Biomarkers for Differential Diagnosis of Sepsis and Other Non-Septic Disease Conditions. Front Cell Infect Microbiol 2022:12, 901433. 10.3389/fcimb.2022.901433.

14. Jeffrey, M., Denny, K.J., Lipman, J. & Conway Morris, A. Differentiating infection, colonisation, and sterile inflammation in critical illness: the emerging role of host-response profiling. Intensive Care Med 2023:49, 760–771. 10.1007/s00134-023-07108-6.

15. Denstaedt, S.J., Singer, B.H. & Standiford, T.J. Sepsis and Nosocomial Infection: Patient Characteristics, Mechanisms, and Modulation. Front Immunol 2018:9, 2446. 10.3389/fimmu.2018.02446.

16. van der Poll, T., Shankar-Hari, M. & Wiersinga, W.J. The immunology of sepsis. Immunity 2021:54, 2450–2464. 10.1016/j.immuni.2021.10.012.

17. Nedeva, C. Inflammation and Cell Death of the Innate and Adaptive Immune System during Sepsis. Biomolecules 2021:11. 10.3390/biom11071011.

18. Wiersinga, W.J. & van der Poll, T. Immunopathophysiology of human sepsis. EBioMedicine 2022:86, 104363. 10.1016/j.ebiom.2022.104363.

19. Dolin, H.H., Papadimos, T.J., Stepkowski, S., Chen, X. & Pan, Z.K. A Novel Combination of Biomarkers to Herald the Onset of Sepsis Prior to the Manifestation of Symptoms. Shock 2018:49, 364–370. 10.1097/SHK.0000000000001010.

20. Laufer, J.M. & Legler, D.F. Beyond migration-Chemokines in lymphocyte priming, differentiation, and modulating effector functions. J Leukoc Biol 2018:104, 301–312. 10.1002/JLB.2MR1217-494R.

21. Doganyigit, Z., Eroglu, E. & Akyuz, E. Inflammatory mediators of cytokines and chemokines in sepsis: From bench to bedside. Hum Exp Toxicol 2022:41, 9603271221078871. 10.1177/09603271221078871.

22. Fox, J.M., Letellier, E., Oliphant, C.J. & Signoret, N. TLR2-dependent pathway of heterologous down-modulation for the CC chemokine receptors 1, 2, and 5 in human blood monocytes. Blood 2011:117, 1851–1860. 10.1182/blood-2010-05-287474.

23. Chaiwut, R. & Kasinrerk, W. Very low concentration of lipopolysaccharide can induce the production of various cytokines and chemokines in human primary monocytes. BMC Res Notes 2022:15, 42. 10.1186/s13104-022-05941-4.

24. Kasprowicz, R., Rand, E., O’Toole, P.J. & Signoret, N. A correlative and quantitative imaging approach enabling characterization of primary cell-cell communication: Case of human CD4(+) T cell-macrophage immunological synapses. Sci Rep 2018:8, 8003. 10.1038/s41598-018-26172-3.

25. Excellence, N.I.f.H.a.C. Sepsis: Recognition, Diagnosis and Early Management [NICE Guideline No.51]. 2016.

26. Yealy, D.M. et al. Recognizing and managing sepsis: what needs to be done? BMC Med 2015:13, 98. 10.1186/s12916-015-0335-2.

27. Fox, J.M., Kasprowicz, R., Hartley, O. & Signoret, N. CCR5 susceptibility to ligand-mediated down-modulation differs between human T lymphocytes and myeloid cells. J Leukoc Biol 2015:98, 59–71. 10.1189/jlb.2A0414-193RR.

28. Team, R.c. R: A Language and Environment for Statistical Computing. In: Computing, R.F.f.S., editor. Vienna, Austria; 2021.

29. Zierk, J. et al. Blood counts in adult and elderly individuals: defining the norms over eight decades of life. Br J Haematol 2020:189, 777–789. 10.1111/bjh.16430.

30. Chen, H. et al. Functional comparison of PBMCs isolated by Cell Preparation Tubes (CPT) vs. Lymphoprep Tubes. BMC Immunol 2020:21, 15. 10.1186/s12865-020-00345-0.

31. Walling, H.W. & Manian, F.A. Predictive Value of Leukocytosis and Neutrophilia for Bloodstream Infection. Infectious Diseases in Clinical Practice 2004:12, 2–6. 10.1097/01.idc.0000104893.16995.0a

32. Zhou, W. et al. Soluble CD14 Subtype in Peripheral Blood is a Biomarker for Early Diagnosis of Sepsis. Lab Med 2020:51, 614–619. 10.1093/labmed/lmaa015.

33. Yang, J. et al. Expansion of a Population of Large Monocytes (Atypical Monocytes) in Peripheral Blood of Patients with Acute Exacerbations of Chronic Obstructive Pulmonary Diseases. Mediators Inflamm 2018:2018, 9031452. 10.1155/2018/9031452.

34. Zhang, D. et al. Frontline Science: COVID-19 infection induces readily detectable morphologic and inflammation-related phenotypic changes in peripheral blood monocytes. J Leukoc Biol 2021:109, 13–22. 10.1002/JLB.4HI0720-470R.

35. Jiang, J. et al. Nonviral infection-related lymphocytopenia for the prediction of adult sepsis and its persistence indicates a higher mortality. Medicine (Baltimore) 2019:98, e16535. 10.1097/MD.0000000000016535.

36. Schaaf, B. et al. Mortality in human sepsis is associated with downregulation of Toll-like receptor 2 and CD14 expression on blood monocytes. Diagn Pathol 2009:4, 12. 10.1186/1746-1596-4-12.

37. Vazquez-Salat, N., Yuhki, N., Beck, T., O’Brien, S.J. & Murphy, W.J. Gene conversion between mammalian CCR2 and CCR5 chemokine receptor genes: a potential mechanism for receptor dimerization. Genomics 2007:90, 213–224. 10.1016/j.ygeno.2007.04.009.

38. Ozanska, A., Szymczak, D. & Rybka, J. Pattern of human monocyte subpopulations in health and disease. Scand J Immunol 2020:92, e12883. 10.1111/sji.12883.

39. Kasten, K.R., Tschop, J., Adediran, S.G., Hildeman, D.A. & Caldwell, C.C. T cells are potent early mediators of the host response to sepsis. Shock 2010:34, 327–336. 10.1097/SHK.0b013e3181e14c2e.

40. Jensen, I.J., Sjaastad, F.V., Griffith, T.S. & Badovinac, V.P. Sepsis-Induced T Cell Immunoparalysis: The Ins and Outs of Impaired T Cell Immunity. J Immunol 2018:200, 1543–1553. 10.4049/jimmunol.1701618.

41. Luperto, M. & Zafrani, L. T cell dysregulation in inflammatory diseases in ICU. Intensive Care Med Exp 2022:10, 43. 10.1186/s40635-022-00471-6.

42. Martin, M.D., Badovinac, V.P. & Griffith, T.S. CD4 T Cell Responses and the Sepsis-Induced Immunoparalysis State. Front Immunol 2020:11, 1364. 10.3389/fimmu.2020.01364.

43. Gao, Y.L. et al. Regulatory T Cells: Angels or Demons in the Pathophysiology of Sepsis? Front Immunol 2022:13, 829210. 10.3389/fimmu.2022.829210.

44. Heidarian, M., Griffith, T.S. & Badovinac, V.P. Sepsis-induced changes in differentiation, maintenance, and function of memory CD8 T cell subsets. Front Immunol 2023:14, 1130009. 10.3389/fimmu.2023.1130009.

45. Trautmann, A. et al. Human CD8 T cells of the peripheral blood contain a low CD8 expressing cytotoxic/effector subpopulation. Immunology 2003:108, 305–312. 10.1046/j.1365-2567.2003.01590.x.

46. Quan, X.Q. et al. Age-related changes in peripheral T-cell subpopulations in elderly individuals: An observational study. Open Life Sci 2023:18, 20220557. 10.1515/biol-2022-0557.

47. Groom, J.R. & Luster, A.D. CXCR3 in T cell function. Exp Cell Res 2011:317, 620–631. 10.1016/j.yexcr.2010.12.017.

48. Contento, R.L. et al. CXCR4-CCR5: a couple modulating T cell functions. Proc Natl Acad Sci U S A 2008:105, 10101–10106. 10.1073/pnas.0804286105.

49. Kmieciak, M. et al. Human T cells express CD25 and Foxp3 upon activation and exhibit effector/memory phenotypes without any regulatory/suppressor function. J Transl Med 2009:7, 89. 10.1186/1479-5876-7-89.

50. Arlettaz, L. et al. CD45 isoform phenotypes of human T cells: CD4(+)CD45RA(-)RO(+) memory T cells re-acquire CD45RA without losing CD45RO. Eur J Immunol 1999:29, 3987–3994. 10.1002/(SICI)1521-4141(199912)29:12<3987::AID-IMMU3987>3.0.CO;2-4.

51. Sutmuller, R.P. et al. Toll-like receptor 2 controls expansion and function of regulatory T cells. J Clin Invest 2006:116, 485–494. 10.1172/JCI25439.

52. Oberg, H.H., Juricke, M., Kabelitz, D. & Wesch, D. Regulation of T cell activation by TLR ligands. Eur J Cell Biol 2011:90, 582–592.10.1016/j.ejcb.2010.11.012.

53. Qin, S. et al. The chemokine receptors CXCR3 and CCR5 mark subsets of T cells associated with certain inflammatory reactions. J Clin Invest 1998:101, 746–754. 10.1172/JCI1422.

54. Balashov, K.E., Rottman, J.B., Weiner, H.L. & Hancock, W.W. CCR5(+) and CXCR3(+) T cells are increased in multiple sclerosis and their ligands MIP-1alpha and IP-10 are expressed in demyelinating brain lesions. Proc Natl Acad Sci U S A 1999:96, 6873–6878. 10.1073/pnas.96.12.6873.

55. Mackay, C.R. CXCR3(+)CCR5(+) T cells and autoimmune diseases: guilty as charged? J Clin Invest 2014:124, 3682–3684. 10.1172/JCI77837.

56. Li, R. et al. Temporary CXCR3 and CCR5 antagonism following vaccination enhances memory CD8 T cell immune responses. Mol Med 2016:22, 497–507. 10.2119/molmed.2015.00218.

57. Baecher-Allan, C., Brown, J.A., Freeman, G.J. & Hafler, D.A. CD4+CD25high regulatory cells in human peripheral blood. J Immunol 2001:167, 1245–1253. 10.4049/jimmunol.167.3.1245.

58. Jonuleit, H. et al. Identification and functional characterization of human CD4(+)CD25(+) T cells with regulatory properties isolated from peripheral blood. J Exp Med 2001:193, 1285–1294. 10.1084/jem.193.11.1285.

59. Dieckmann, D., Bruett, C.H., Ploettner, H., Lutz, M.B. & Schuler, G. Human CD4(+)CD25(+) regulatory, contact-dependent T cells induce interleukin 10-producing, contact-independent type 1-like regulatory T cells [corrected]. J Exp Med 2002:196, 247–253. 10.1084/jem.20020642.

60. Komai-Koma, M., Jones, L., Ogg, G.S., Xu, D. & Liew, F.Y. TLR2 is expressed on activated T cells as a costimulatory receptor. Proc Natl Acad Sci U S A 2004:101, 3029–3034. 10.1073/pnas.0400171101.

61. Vincent, J.L., Opal, S.M., Marshall, J.C. & Tracey, K.J. Sepsis definitions: time for change. Lancet 2013:381, 774–775. 10.1016/S0140-6736(12)61815-7.

62. Ma, L. et al. Role of interleukin-6 to differentiate sepsis from non-infectious systemic inflammatory response syndrome. Cytokine 2016:88, 126–135.10.1016/j.cyto.2016.08.033.

63. Metzemaekers, M., Vanheule, V., Janssens, R., Struyf, S. & Proost, P. Overview of the Mechanisms that May Contribute to the Non-Redundant Activities of Interferon-Inducible CXC Chemokine Receptor 3 Ligands. Front Immunol 2017:8, 1970. 10.3389/fimmu.2017.01970.

64. Wilson, J.C. et al. Integrated miRNA/cytokine/chemokine profiling reveals severity-associated step changes and principal correlates of fatality in COVID-19. iScience 2022:25, 103672. 10.1016/j.isci.2021.103672.

65. Coperchini, F., Chiovato, L. & Rotondi, M. Interleukin-6, CXCL10 and Infiltrating Macrophages in COVID-19-Related Cytokine Storm: Not One for All But All for One! Front Immunol 2021:12, 668507. 10.3389/fimmu.2021.668507.

66. Hsu, R.J. et al. The Role of Cytokines and Chemokines in Severe Acute Respiratory Syndrome Coronavirus 2 Infections. Front Immunol 2022:13, 832394. 10.3389/fimmu.2022.832394.

67. Xue, M. et al. Early and dynamic alterations of Th2/Th1 in previously immunocompetent patients with community-acquired severe sepsis: a prospective observational study. J Transl Med 2019:17, 57. 10.1186/s12967-019-1811-9.

68. Liu, Y., Wang, X. & Yu, L. Th17, rather than Th1 cell proportion, is closely correlated with elevated disease severity, higher inflammation level, and worse prognosis in sepsis patients. J Clin Lab Anal 2021:35, e23753. 10.1002/jcla.23753.

69. Costa, R.T. et al. T helper type cytokines in sepsis: time-shared variance and correlation with organ dysfunction and hospital mortality. Braz J Infect Dis 2019:23, 79–85. 10.1016/j.bjid.2019.04.008.

70. Zanza, C. et al. Cellular Immuno-Profile in Septic Human Host: A Scoping Review. Biology (Basel) 2022:11. 10.3390/biology11111626.

71. Stewart, J.C., Villasmil, M.L. & Frampton, M.W. Changes in fluorescence intensity of selected leukocyte surface markers following fixation. Cytometry A 2007:71, 379–385. 10.1002/cyto.a.20392.

72. Sakkestad, S.T., Skavland, J. & Hanevik, K. Whole blood preservation methods alter chemokine receptor detection in mass cytometry experiments. J Immunol Methods 2020:476, 112673. 10.1016/j.jim.2019.112673.

73. Capelle, C.M. et al. Standard Peripheral Blood Mononuclear Cell Cryopreservation Selectively Decreases Detection of Nine Clinically Relevant T Cell Markers. Immunohorizons 2021:5, 711–720. 10.4049/immunohorizons.2100049.

74. Cabrera-Perez, J., Condotta, S.A., Badovinac, V.P. & Griffith, T.S. Impact of sepsis on CD4 T cell immunity. J Leukoc Biol 2014:96, 767–777. 10.1189/jlb.5MR0114-067R.

75. Flaishon, L. et al. Expression of the chemokine receptor CCR2 on immature B cells negatively regulates their cytoskeletal rearrangement and migration. Blood 2004:104, 933–941. 10.1182/blood-2003-11-4013.

76. Zhang, C. et al. B-Cell Compartmental Features and Molecular Basis for Therapy in Autoimmune Disease. Neurol Neuroimmunol Neuroinflamm 2021:8. 10.1212/NXI.0000000000001070.

77. Castanheira, F. et al. CCR5-Positive Inflammatory Monocytes are Crucial for Control of Sepsis. Shock 2019:52, e100–e106. 10.1097/SHK.0000000000001301.

78. Olive, A.J., Gondek, D.C. & Starnbach, M.N. CXCR3 and CCR5 are both required for T cell-mediated protection against C. trachomatis infection in the murine genital mucosa. Mucosal Immunol 2011:4, 208–216. 10.1038/mi.2010.58.

79. Colvin, R.A., Campanella, G.S., Sun, J. & Luster, A.D. Intracellular domains of CXCR3 that mediate CXCL9, CXCL10, and CXCL11 function. J Biol Chem 2004:279, 30219–30227. 10.1074/jbc.M403595200.

80. Karin, N., Wildbaum, G. & Thelen, M. Biased signaling pathways via CXCR3 control the development and function of CD4+ T cell subsets. J Leukoc Biol 2016:99, 857–862. 10.1189/jlb.2MR0915-441R.

81. Rabin, R.L. et al. CXCR3 is induced early on the pathway of CD4+ T cell differentiation and bridges central and peripheral functions. J Immunol 2003:171, 2812–2824. 10.4049/jimmunol.171.6.2812.

82. Hu, J.K., Kagari, T., Clingan, J.M. & Matloubian, M. Expression of chemokine receptor CXCR3 on T cells affects the balance between effector and memory CD8 T-cell generation. Proc Natl Acad Sci U S A 2011:108, E118–127. 10.1073/pnas.1101881108.

83. Meiser, A. et al. The chemokine receptor CXCR3 is degraded following internalization and is replenished at the cell surface by de novo synthesis of receptor. J Immunol 2008:180, 6713–6724. 10.4049/jimmunol.180.10.6713.

84. O’Boyle, G. et al. Chemokine receptor CXCR3 agonist prevents human T-cell migration in a humanized model of arthritic inflammation. Proc Natl Acad Sci U S A 2012:109, 4598–4603. 10.1073/pnas.1118104109.

85. Hoerning, A. et al. Subsets of human CD4(+) regulatory T cells express the peripheral homing receptor CXCR3. Eur J Immunol 2011:41, 2291–2302. 10.1002/eji.201041095.

86. Burton, R.J. et al. Conventional and unconventional T cell responses contribute to the prediction of clinical outcome and causative bacterial pathogen in sepsis patients. Clin Exp Immunol 2024. 10.1093/cei/uxae019.

87. Chami, B. et al. CXCR3 plays a critical role for host protection against Salmonellosis. Sci Rep 2017:7, 10181. 10.1038/s41598-017-09150-z.

88. Griffith, J.W., Sokol, C.L. & Luster, A.D. Chemokines and chemokine receptors: positioning cells for host defense and immunity. Annu Rev Immunol 2014:32, 659–702. 10.1146/annurev-immunol-032713-120145.

