## supplemental material for "Blood immune profiles reveal a CXCR3/CCR5 axis of dysregulation in early sepsis"

### 1- supplemental figures and tables legends:

S1: Antibody panels used for blood cell phenotyping. The blood cell panel (BCP) and T cell activation panel (TCAP). Includes antibody names, source and manufacturer references. The matrixes indicate the antibody combinations used for full staining (BCP or TCAP) plus Fluorescence minus one (FMO) controls.

S2: General flow cytometry gating strategy for analysis. A) For general cell types within blood isolated PBMCs; B) BCP-specific: to select monocytes, T ad B cells according to CD14, CD3 and CD19 expression, respectively before analysing the percentage of cells expressing the indicated surface markers; C) TCAP-specific: to separate $CD8^+$ and $CD4^+$ from lymphocytes before analysing the percentage of cells expressing the indicated markers.

S3: Monocytes subsets. A) Scatter plot showing percentage of $CD14^+$ monocytes for the healthy controls and suspected sepsis groups, with Mean values +/-SD; B) Flow cytometry scatter plot showing CD14/CD16 gating on classical, intermediate and non-classical monocytes (CM, IM and NCM, respectively) and graph plotting the percentage of monocytes for each subpopulation, with Mean +/-SD.

S4: Expression of CCR1, CCR7 and TLR2 on monocytes subpopulations. Boxplots showing the percentage of positive cells for each marker, Boxplots for both healthy (white) and suspected sepsis (grey) groups are shown with all points plotted. Mann-Whitney tests showed no statistical differences.

S5: PCA for BCP panel variables: Principal component analysis (PCA) scores plot for the first two components for the blood cell panel dataset with healthy controls in black and sepsis-suspected patients in red. The biplot shows the loadings as vectors (black arrows) and scores by sample names in grey.

S6: Flow cytometry gating strategy for subpopulations with TCAP. A) gating for single marker analysis, B) CXCR3/CCR5 co-expression, C) CD45RO/CD25 double positive cells, all with gates set based on the relevant FMO controls.

S7: Markers expression profiles on T cells subpopulations. Frequency of $CD8^{+(high)}$ and $CD4^+$ T cells that are A) CXCR3/CCR5 double positive, B) Memory T cells ($CD45RO^+$), C) CXCR3/CCR5 double positive Type-1 like regulatory T cells (CD45RO/CD25), D) TLR2 positive memory T cells ($CD45RO^+$). Boxplots for both healthy and suspected sepsis groups are shown with all points plotted. Statistical significance (adjusted p values) were determined using Mann-Whitney tests with adjustment for multiple comparisons using Benjamini, Krieger and Yekutiel secondary test.

S8: PCA for all TCAP variables. Principal component analysis (PCA) scores plot for the first two components for the T cell activation panel dataset with healthy controls in black and sepsis-suspected patients in red. The biplot shows the loadings as vectors (black arrows) and scores by sample names in grey.

S9: Spearman correlation analysis for CC and sCD14 profiles on sepsis subgroups. Heatmaps showing correlation matrix after hierarchical clustering for confirmed and unconfirmed sepsis subgroups.

S10: PCA graphs identifying sepsis subgroups. Principal component analysis plots for the first two components grouping healthy controls (black), confirmed sepsis (sepsis, red) and unconfirmed sepsis (purple) cases for variables from (A) the three separate datasets and (B) the three datasets combined. The scores plot obtained using variables from all datasets (see Table S2) shows grouping of healthy controls (black ellipse) but does not separate the patient sub-groups.

Table S1: relevant clinical information regarding patients recruited upon admission to York hospital ED with suspected sepsis, including microbiology tests results and clinical diagnostic upon discharge.

Table S2: Full dataset of variables and measures incorporated in the study analysis.

### 2- Supplemental Figures

#### Figure S1:

| **Antibodies** | |
|---|---|
| **sources** | **Ref number** |
| Biolegends | 301038 |
| Biolegends | 302048 |
| In house | Clone MC5 |
| Biolegends | 309708 |
| Biolegends | 353236 |
| Biolegends | 302238 |
| Biolegends | 362908 |
| Biolegends | 325632 |
| Biolegends | 300470 |

Blood cell panel (BCP)

| | FMO BV421 | FMO BV510 | FMO A488 | FMO PE | FMO PE/Dazzle 594 | FMO BV650 | FMO APC | FMO PerCP | FMO APC/Fire 750 | FMO 808 | BCP | Unstained |
|---|---|---|---|---|---|---|---|---|---|---|---|---|
| BV421 CD16 | | x | x | x | x | x | x | x | x | x | x | |
| BV510 CCR2 | x | | x | x | x | x | x | x | x | x | x | |
| A488 CCR5 | x | x | | x | x | x | x | x | x | x | x | |
| PE TLR2.1 | x | x | x | | x | x | x | x | x | x | x | |
| PE/Dazzle 594 CCR7 | x | x | x | x | | x | x | x | x | x | x | |
| BV650 CD19 | x | x | x | x | x | | x | x | x | x | x | |
| APC CCR1 | x | x | x | x | x | x | | x | x | x | x | |
| PerCP CD14 | x | x | x | x | x | x | x | | x | x | x | |
| APC/Fire 750 CD3 | x | x | x | x | x | x | x | x | | x | x | |
| 808 Live/Dead | x | x | x | x | x | x | x | x | x | | x | |

x = includes this fluorophore

| **Antibodies** | |
|---|---|
| **sources** | **Ref number** |
| Biolegends | 302630 |
| Abcam | ab16894 |
| Biolegends | 353236 |
| Biolegends | 300548 |
| In house | Clone MC5 |
| Biolegends | 304218 |
| Biolegends | 344746 |

T-cell activation panel (TCAP)

| | FMO BV421 | FMO FITC | FMO PE | FMO PE/Dazzle 594 | FMO A647 | FMO A700 | FMO APC/Fire 750 | FMO 808 | TCAP | Unstained |
|---|---|---|---|---|---|---|---|---|---|---|
| BV421 CD25 | | x | x | x | x | x | x | x | x | |
| FITC TLR2.5 | x | | x | x | x | x | x | x | x | |
| PE TLR2.1 CXCR3 | x | x | | x | x | x | x | x | x | |
| PE/Dazzle 594 CD4 | x | x | x | | x | x | x | x | x | |
| A647 CCR5 | x | x | x | x | | x | x | x | x | |
| A700 CD45RO | x | x | x | x | x | | x | x | x | |
| APC/Fire 750 CD8 | x | x | x | x | x | x | | x | x | |
| 808 Live/Dead | x | x | x | x | x | x | x | | x | |

x = includes this fluorophore

#### Figure S2:

**A.** General Blood-isolated cells gating:

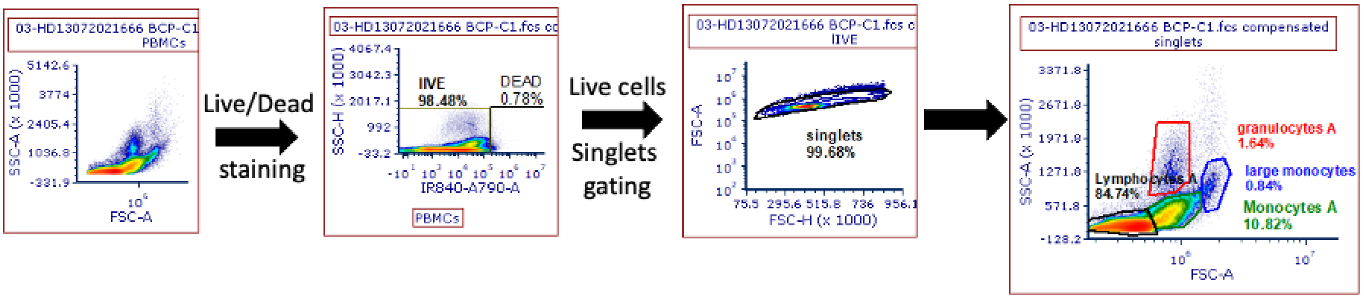


**B.** Blood cell Panel (BCP) specific gating:

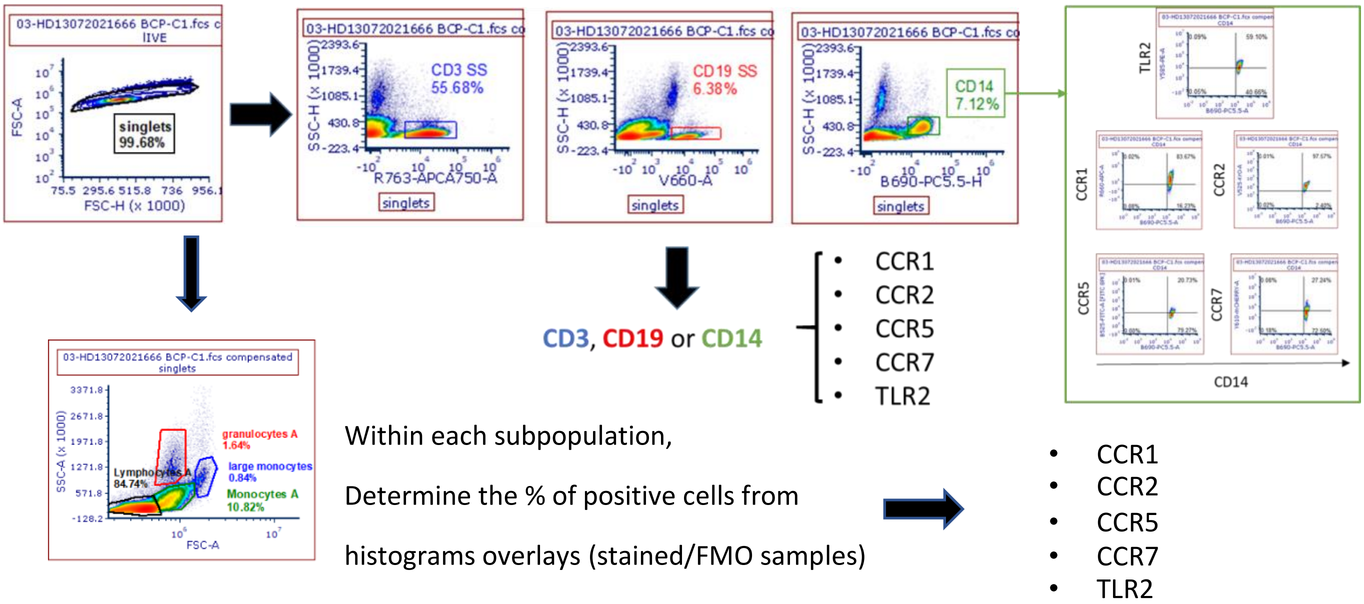


**C.** T cell Panel (TCAP) specific gating strategy:

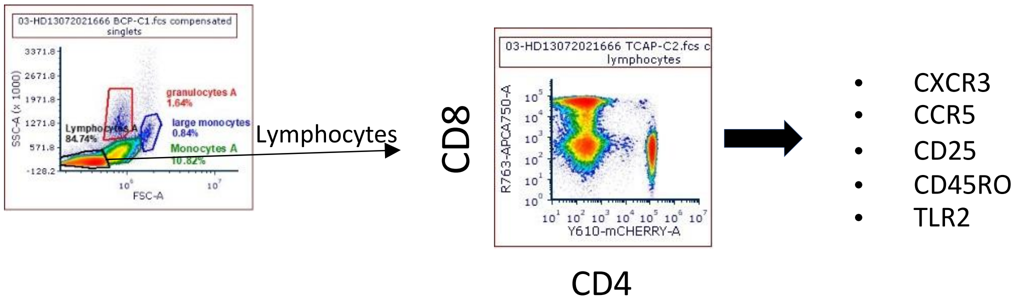

**Figure S3:**

**A.** CD14$^+$ cells in monocytes population

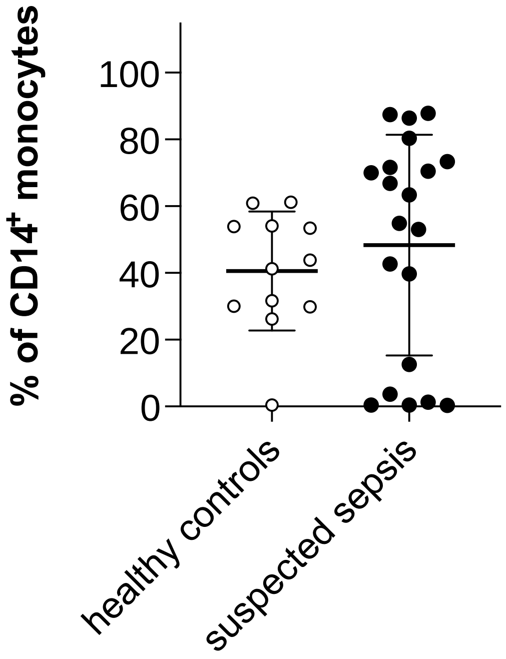


**B.** CD14 and CD16 expression to define Classical, Intermediate and non-classical monocyte subsets (CM, IM and NCM)

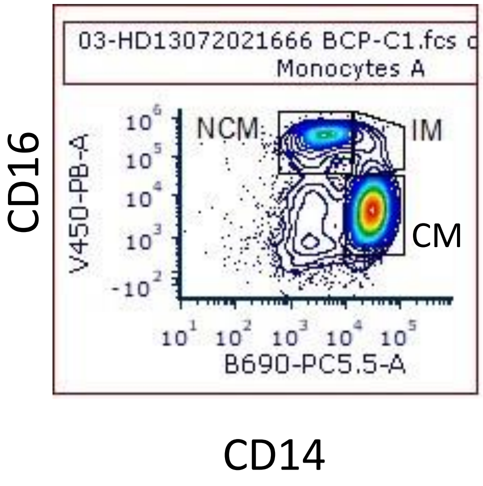


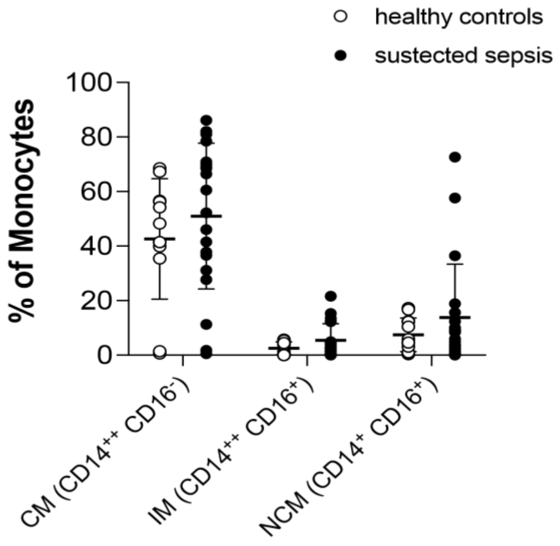

**Figure S4:**

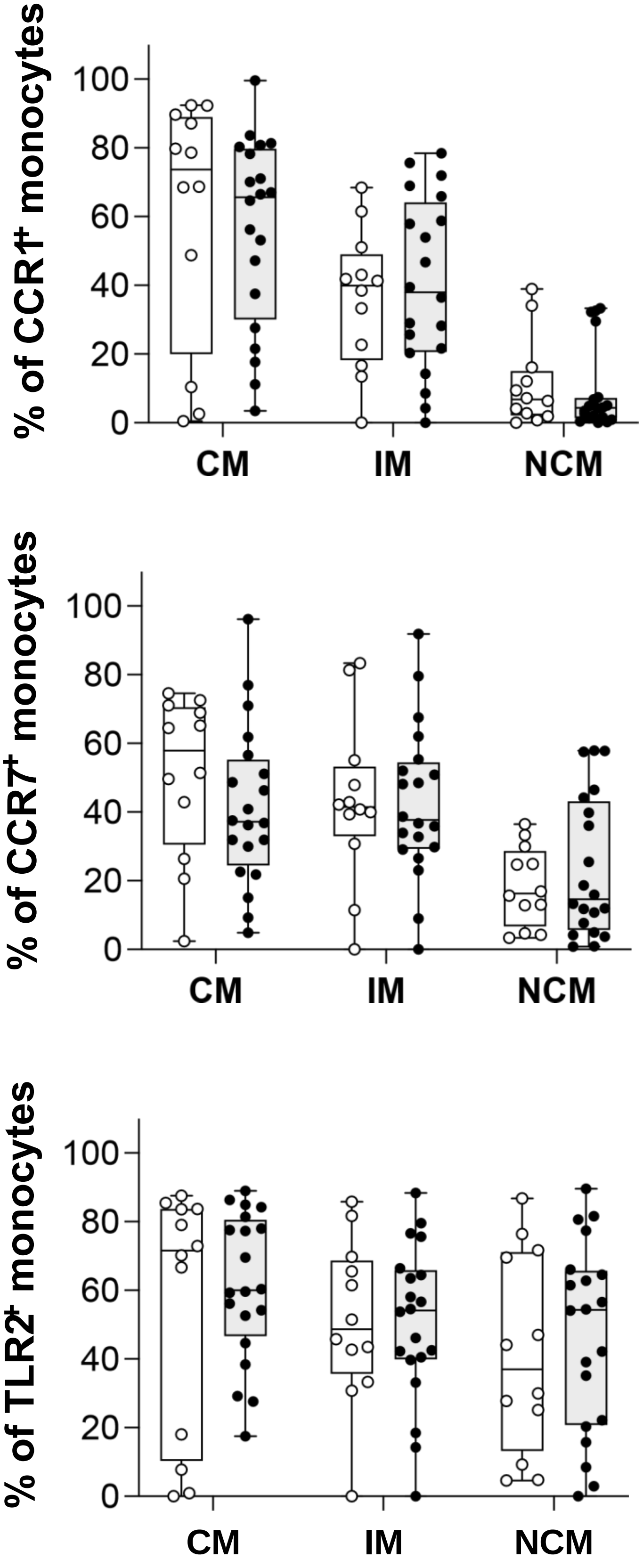

% of CCR1+ monocytes
100
80
60
40
20
0
CM
IM
NCM
% of CCR7+ monocytes
100
80
60
40
20
0
CM
IM
NCM
% of TLR2+ monocytes
100
80
60
40
20
0
CM
IM
NCM

**Figure S5:**

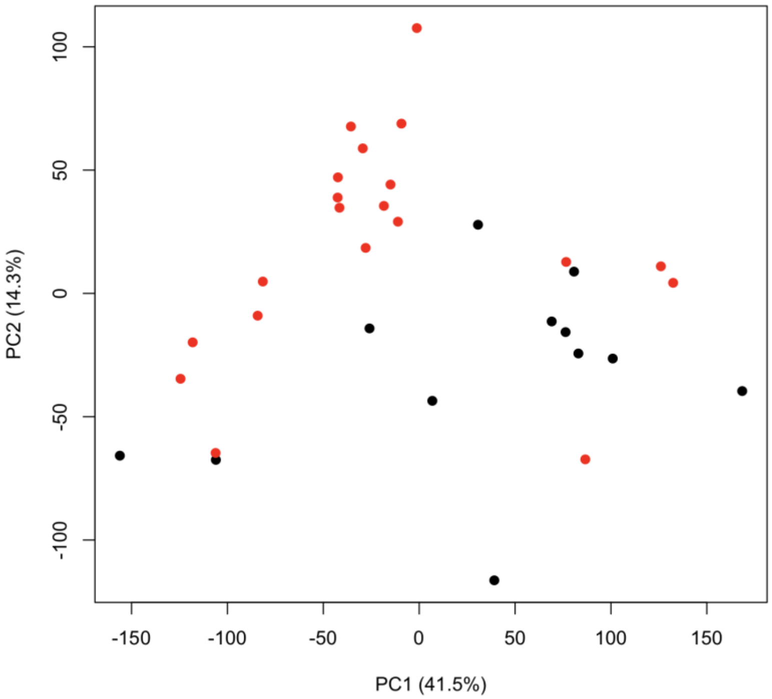
PC2 (14.3%)
PC1 (41.5%)

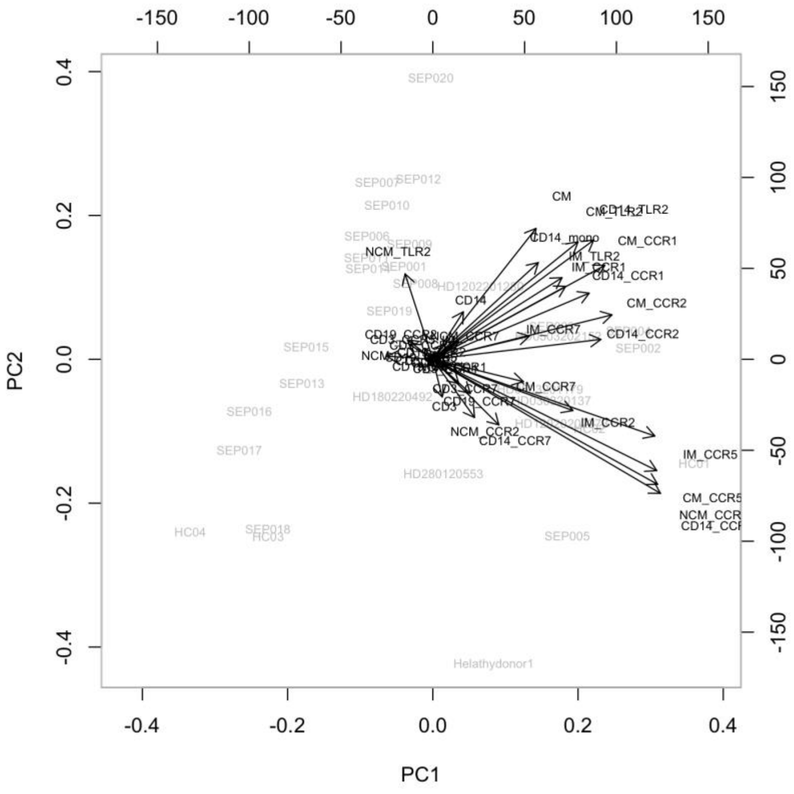
PC1
PC2
SEP020
SEP012
SEP007
SEP010
SEP015
SEP013
SEP016
SEP017
HC04
SEP018
HC03
SEP005
HD280120553
HD180220492
SEP002
Helathydonor1
CM
CM_TLR2
CD14_TLR2
CD14_mono
CM_CCR1
IM_TLR2
CD14_CCR1
CM_CCR2
CD14_CCR2
CD14
NCM_TLR2
IM_CCR7
CM_CCR7
IM_CCR2
NCM_CCR2
CD14_CCR7
CD19_CCR7
IM_CCR5
CM_CCR5

### Figure S6:

**A.**

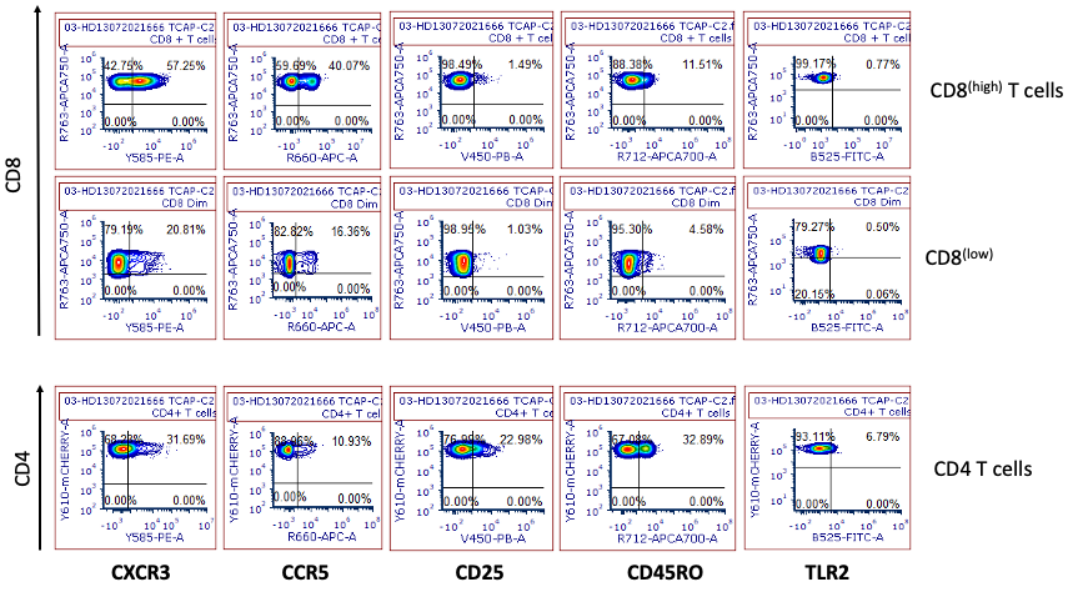


**B.**

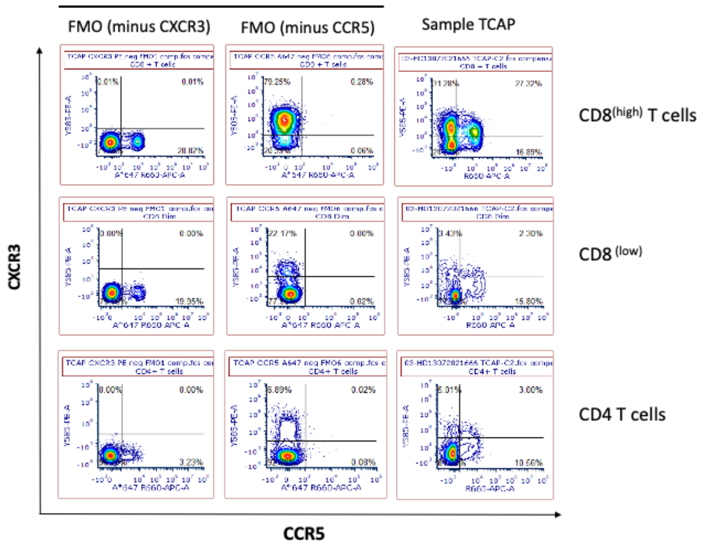


**C.**

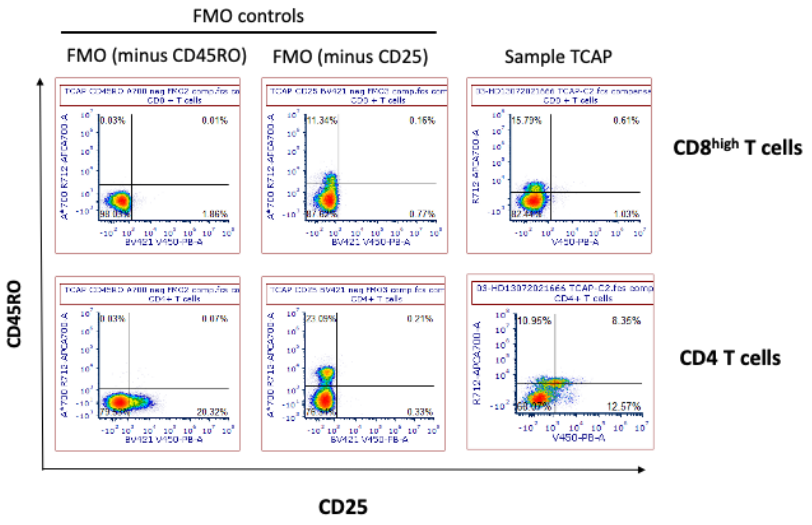

**Figure S7:**

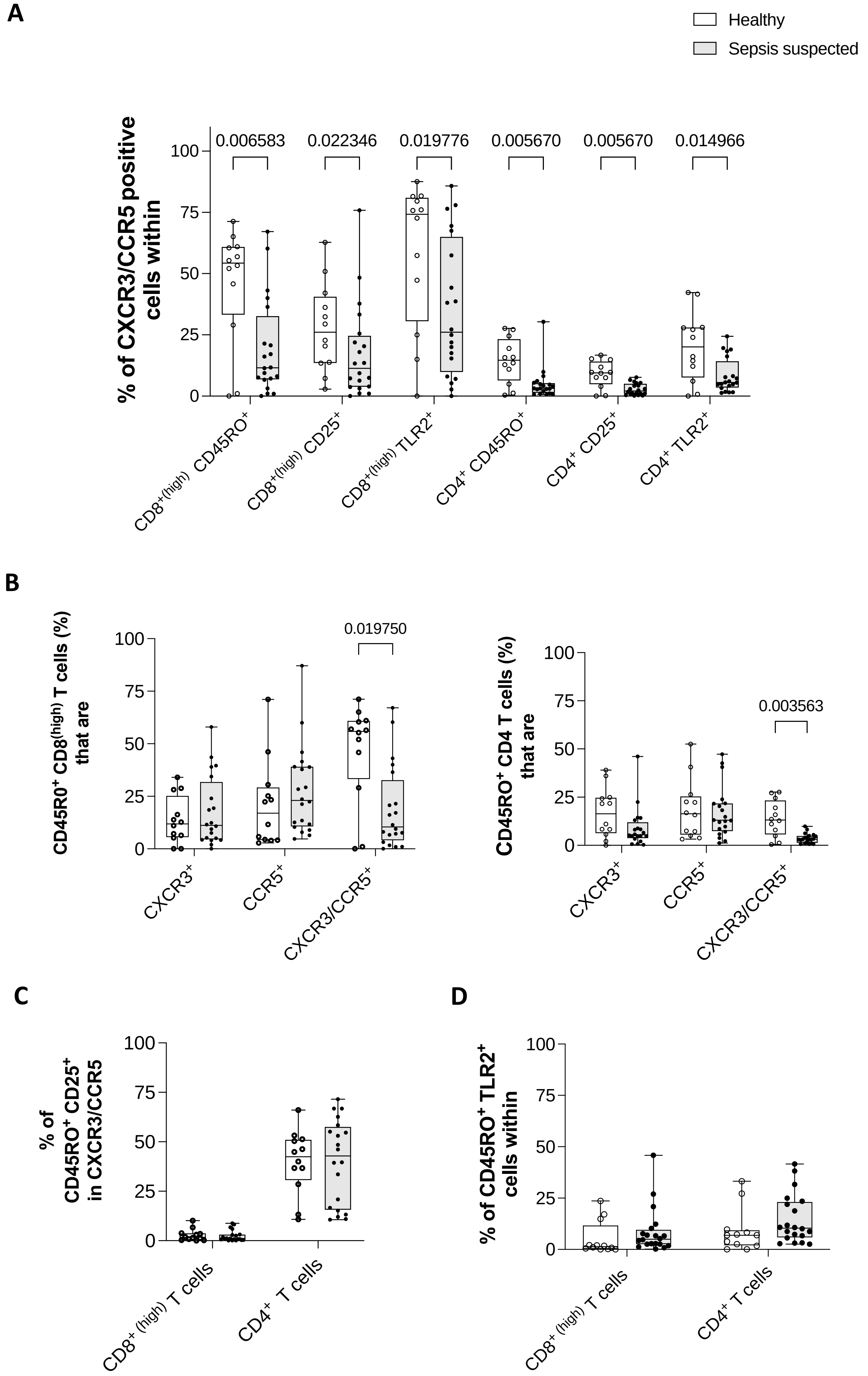
A
Healthy
Sepsis suspected
% of CXCR3/CCR5 positive cells within
0.006583
0.022346
0.019776
0.005670
0.005670
0.014966
100
75
50
25
0
CD8+(high) CD45RO+
CD8+(high) CD25+
CD8+(high) TLR2+
CD4+ CD45RO+
CD4+ CD25+
CD4+ TLR2+
B
CD45R0+ CD8(high) T cells (%) that are
0.019750
CXCR3+
CCR5+
CXCR3/CCR5+
CD45RO+ CD4 T cells (%) that are
0.003563
C
% of CD45RO+ CD25+ in CXCR3/CCR5
CD8+ (high) T cells
CD4+ T cells
D
% of CD45RO+ TLR2+ cells within

**Figure S8:**

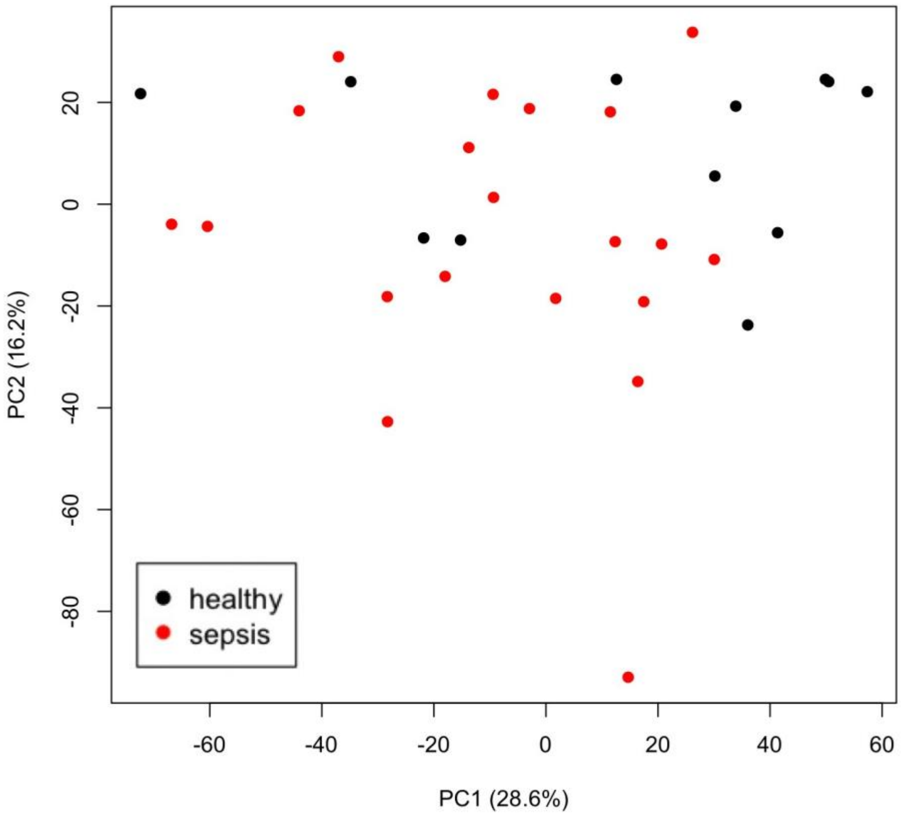
healthy
sepsis
PC1 (28.6%)
PC2 (16.2%)

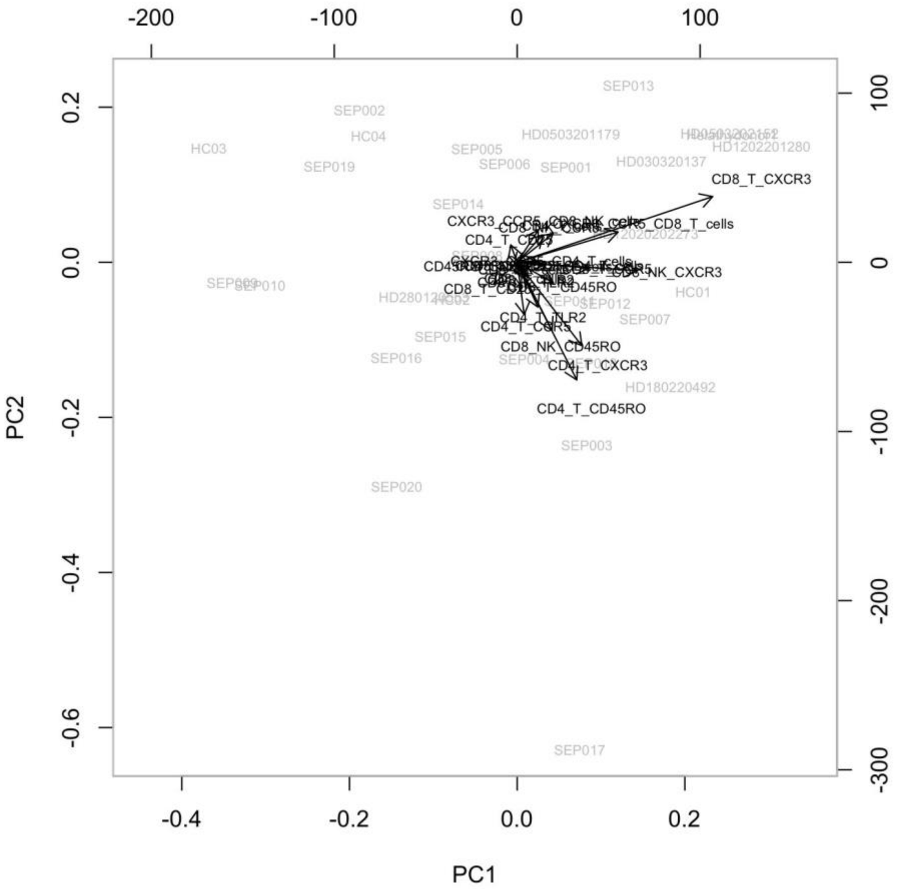
PC1
PC2
CD8_T_CXCR3
CD4_T_CD45RO
CD4_T_CXCR3
CD8_NK_CD45RO
SEP013
SEP002
SEP020
SEP017
SEP003

#### Figure S9:

### Confirmed Sepsis

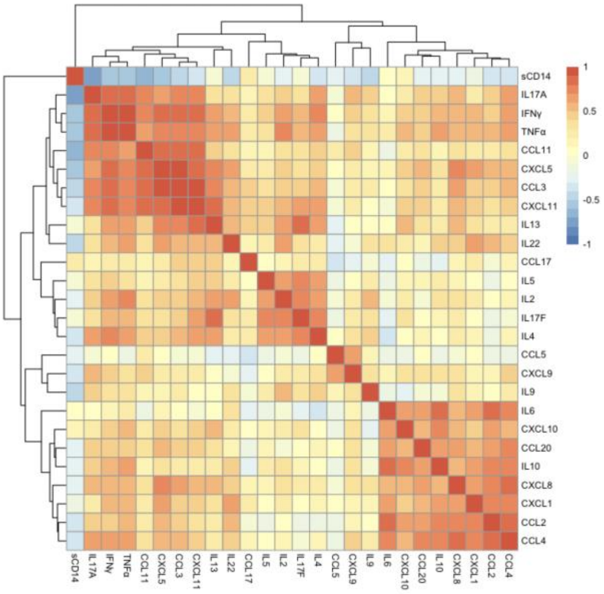


### Unconfirmed Sepsis

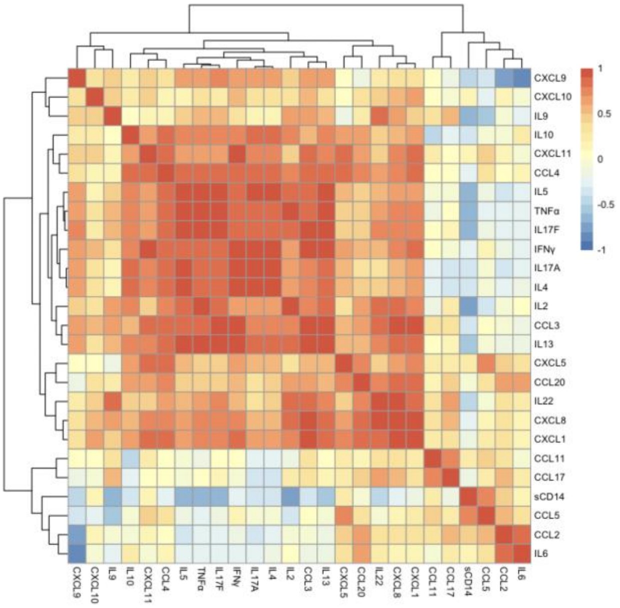

### Figure S10:

**A.**

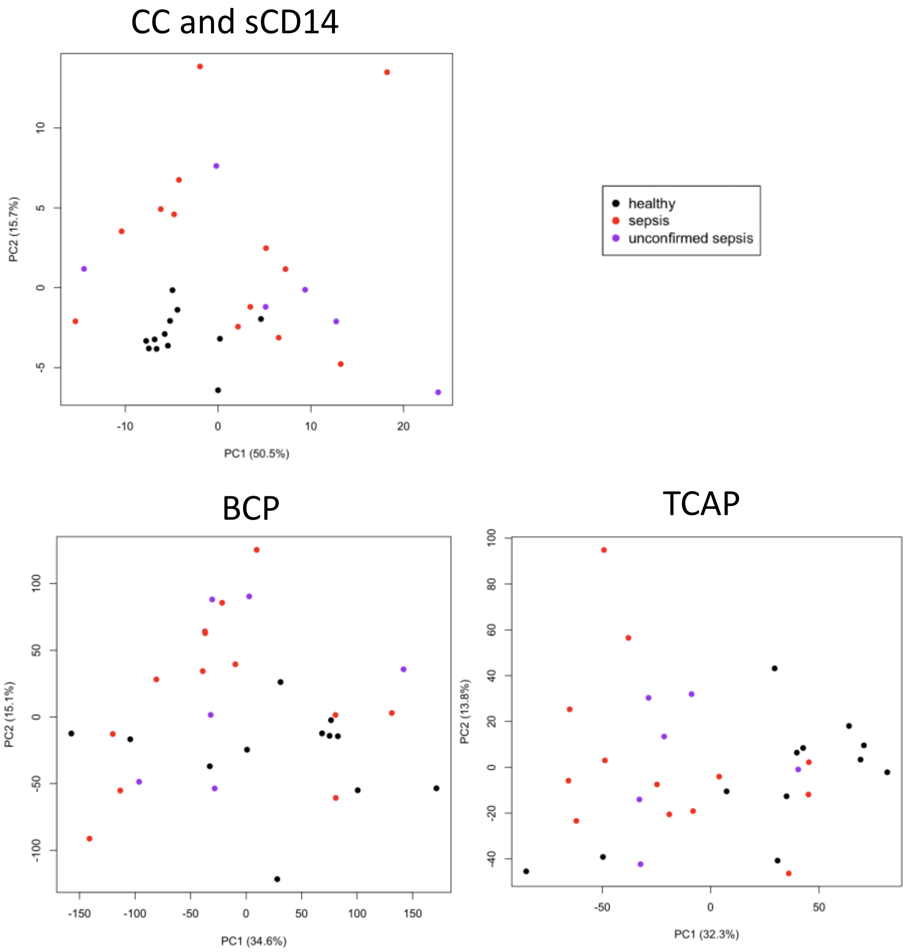


**B.**

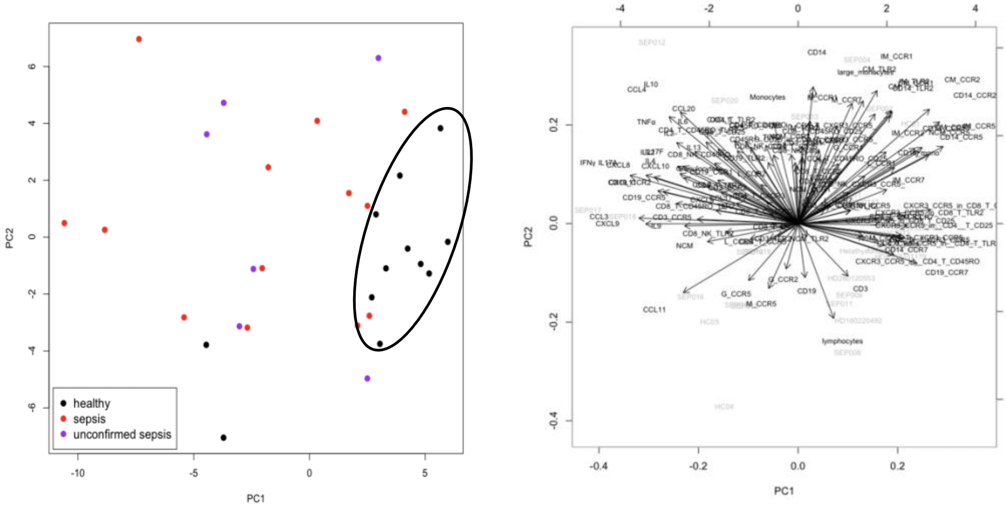

### 3- Supplemental Tables:

#### Table S1:

| Patient | Age | Sex | Presenting Complaint | Diagnosis in ED | WBC upon enrollement (109/L) normal range 4.5-11 | Length of Stay | Status at Discharge | Vasoactive Infusion | Culture Results | CXR Results | Peak CRP within 48 hours | Gram Positive Infection? | Diagnosis on Discharge | Sepsis status on Discharge |
|---|---|---|---|---|---|---|---|---|---|---|---|---|---|---|
| SEP001 | 37 | Female | Drowsiness | Chest sepsis | 24.5 | 2 days | Alive | No | 2x BCs Negat | Right basal consolidation | 152 | Possible - CAP but no positive cultures | CAP | Confirmed |
| SEP002 | 60 | Male | Confusion | Sepsis 2y to cellulitis | 18.8 | 16 days | Alive | No | 2x BCs Negat | N/A | 164 | Possible - swab grew S aureus | Sepsis of unknown origin, AKI, hypokalaemia, hyponatraemia, venous leg ulcers | Confirmed |
| SEP003 | 56 | Female | Shortness of breath | Chest sepsis | 17.7 | 77 days | Alive | Yes | 2x BCs Negat | Bilateral basal haziness | 165 | Unlikely - ? Viral exacerbation of asthma. | Infective exacerbation of asthma, metapneumpvirus positibe, AKI, C. difficicle infection, UTI | Confirmed |
| SEP004 | 54 | Male | SOB/cough | CAP | 21.7 | 1 day | Alive | No | 2x BCs Nega | Slight haziness right side | 137 | Unlikely | IECOPD | Unconfirmed |
| SEP005 | 91 | Female | SOB, confusion | Pneumonia | 5.7 | Less than 24 h. | **Dead** | No | 2x BCs Negat | Bilateral consolidation | 156 | Possible - severe CAP but no positive cultures | CAP | Confirmed |
| SEP006 | 93 | Male | SOB | No diagnosis stated | 10.8 | 1 day | Alive | No | 2x BCs Negat | New consolidation | 17 | Unlikely - CAP but no positive cultures and low CRP | CAP | Confirmed |
| SEP007 | 82 | Male | SOB | CAP | 9.1 | 1 day | Alive | No | 2x BCs Negat | Nil acute | 6 | Unlikely - CXR clear and CRP very low | CAP | Unconfirmed |
| SEP008 | 84 | Female | SOB | Possible sepsis, possible l | 8.1 | 7 days | Alive | No | 2x BCs Negat | No consolidation, heart failure | 6 | Unlikely - CXR heart failure and CRP very low | Heart failure | Unconfirmed |
| SEP009 | 87 | Female | SOB | Cellulitis | 15.4 | 3 days | Alive | **withdrew from study on the 28/05/2021** | | | | | | |
| SEP010 | 78 | Female | SOB/rigors | cellulitis | 20.6 | 4 days | Alive | No | BCs - Serratia | N/A | 149 | Possible - S aureus and cellulits and responded to flucloxacillin | Cellulitis | Confirmed |
| SEP011 | 70 | Female | SOB | LRTI, Supraglotitis | 6.5 | 1 day | Alive | No | Culture resu | N/A | less than 1 | Unlikely - low CRP | Laryngopharyngeal Reflux | Confirmed |
| SEP012 | 55 | Male | SOB | Chest sepsis | 27.7 | 6 days | Alive | No | 2x BCs Negat | Bilateral changes consistent with bronchiectasis | 13 | Unlikely - bronchiectasis with Pseudomonas in sputum | Infective exacerbation of his bronchiectasis | Unconfirmed |
| SEP013 | 82 | Female | SOB | Acute infection-source u | 10.9 | 9 days | Alive | No | 2x BCs Negat | Bilateral bronchiectasis changes, suspected post COVID changes | 33 | Unlikely - treated as pulmonary fibrosis | Pulmonary fibrosis | Unconfirmed |
| SEP014 | 79 | Male | SOB/cough | Possible PE, possible pne | 14.8 | 7 days | Alive | No | 2x BCs Negat | Subtle basal changes, possible aspiration pneumonia | 32 | Unlikely - H influenzae in sputum most likely pathogen | Aspiration pneumonia | Confirmed |
| SEP015 | 60 | Male | SOB | Chest sepsis | 12.4 | 3 days | Alive | No | 2x BCs Negat | Nil acute | 3 | Unlikely - antibiotics stopped on post take ward round | Stridor caused by bilateral vocal cord palsy | Confirmed |
| SEP016 | 101 | Male | Confusion/cough | CAP | 7.9 | 4 days | Alive | No | 2x BCs Nega | Left lower zone consolidation | 8 | Possible - CAP but no positive cultures and low CRP | CAP | Confirmed |
| SEP017 | 78 | Male | Confusion | LRTI | 6.8 | 5 days | Alive | No | 2x BCs posit | Nil acute | 134 | No - proven gram negative bacteraemia from urine infection. | Sepsis-no source documented | Confirmed |
| SEP018 | 75 | Male | SOB/drowsy | IECOPD | 15 | 2 days | Alive | No | 2x BCs posit | Right lower zone, suscped consolidation | 5 | Unlikely- low CRP and previous H influenzae in sputum | CAP | Confirmed |
| SEP019 | 21 | Female | SOB | Chest sepsis | 2.8 | 6 days | Alive | No | 2x BCs Nega | Bilateral changes, suspected COVID Pneumonia (NB test positive f | 41 | Unlikely - low CRP and CXR changes typical for COVID | COVID-19 | Unconfirmed |
| SEP020 | 63 | Male | SOB | COPD | 15.5 | 23 days | **Dead** | No | 2x BCs, 1 bo | Bilateral fibrosis, suscpected superadded infective changes | 152 | Possible - but no positive cultures | Pneumonia | Confirmed |

SOB: Shortness of breath
CAP: Community-acquired pneumonia
LRTI: Lower respiratory tract infection
PE: Pulmonary embolism
IECOPD: Infective exacerbation of chronic obstructive pulmonary disease
AKI: Acute kidney injury
UTI: Urinary tract infection

#### Table S2:

| | 1- CC panel Log2 values | | | | | | | | | | | | | | | | | | | | | | | | | 2- sCD14 ELISA (µg/ml) |
|---|---|---|---|---|---|---|---|---|---|---|---|---|---|---|---|---|---|---|---|---|---|---|---|---|---|---|
| Individuals | CXCL8 | CXCL10 | CCL11 | CCL17 | CCL2 | CCL5 | CCL3 | CXCL9 | CXCL5 | CCL20 | CXCL1 | CXCL11 | CCL4 | IL-5 | IL-13 | IL-2 | IL-6 | IL-9 | IL-10 | IFN-γ | TNF-α | IL-17A | IL-17F | IL-4 | IL-22 | sCD14 |
| Helathy donor 1 | 2.158499 | 6.815704 | 6.836783 | 5.358431 | 8.939374 | 9.985016 | 2.65764 | 1.9855 | 6.627972 | 2.094236 | 5.31034 | 1.918386 | 0.111031 | 3.012569 | 1.613532 | 1.147307 | 4.019257 | -0.152 | 1.24184 | 3.210233 | 0.687061 | -2.32193 | -0.21759 | 3.811471 | 4.693208 | 1.0391 |
| HD280120553 | 1.914565 | 5.923648 | 5.363381 | 5.435129 | 8.717471 | 10.07988 | 2.65764 | 1.9855 | 5.238978 | 2.094236 | 5.31034 | 1.918386 | 0.111031 | 3.546586 | 5.568944 | 0.389567 | 2.234195 | -0.152 | 1.843984 | 1.166715 | 1.014355 | -1.55427 | -0.21759 | 2.799087 | 0.007196 | 0.72727 |
| HD1202201280 | 3.008092 | 5.28762 | 4.499463 | 5.218587 | 8.045296 | 11.37542 | 2.65764 | 1.9855 | 7.442944 | 2.094236 | 5.31034 | 1.918386 | 0.111031 | 0.992768 | 1.613532 | 1.871844 | -0.152 | -0.152 | -0.12816 | 0.687061 | 0.687061 | -2.32193 | -0.21759 | 1.166715 | 0.650765 | 0.25845 |
| HD12020202273 | 3.426131 | 6.319853 | 6.318534 | 6.851749 | 9.187748 | 9.053885 | 2.65764 | 1.9855 | 6.893969 | 2.094236 | 5.31034 | 1.918386 | 0.111031 | 0.063503 | 1.613532 | 0.827819 | -0.57777 | -0.152 | 0.169925 | 0.687061 | 1.087463 | -2.32193 | 1.922198 | 1.627607 | 1.82985 | 1.55738 |
| HD180220492 | 1.914565 | 6.046469 | 5.987548 | 5.746985 | 8.860466 | 10.71425 | 4.744699 | 1.9855 | 5.8591 | 3.531693 | 6.798828 | 1.918386 | 0.111031 | 1.114367 | 1.613532 | -0.02915 | 0.327687 | -0.03653 | 0.541019 | 1.411426 | 1.192194 | -0.9173 | 1.176323 | 2.496974 | 0.550901 | 0.51652 |
| HD030320137 | 3.185787 | 5.741952 | 6.225053 | 6.20506 | 8.4798 | 13.6479 | 3.736064 | 1.9855 | 9.423767 | 2.094236 | 7.802969 | 4.56581 | 0.111031 | 2.693766 | 6.40446 | 0.589763 | 1.835924 | 1.92391 | 1.235727 | 3.902556 | 4.523562 | 0.849599 | 2.428946 | 3.33985 | 0.996389 | 0.91006 |
| HD0503201179 | 1.914565 | 6.196843 | 5.915879 | 4.807097 | 8.83145 | 10.44535 | 2.65764 | 1.9855 | 5.556123 | 2.094236 | 5.31034 | 1.918386 | 0.111031 | 2.800123 | 1.613532 | -0.08927 | -0.02915 | -0.152 | 1.214125 | 0.687061 | 0.687061 | -2.32193 | -0.21759 | 2.017922 | -0.97143 | 1.15092 |
| HD0503202152 | 1.914565 | 4.82985 | 6.20637 | 5.354029 | 8.338313 | 10.19994 | 2.65764 | 1.9855 | 5.850374 | 2.094236 | 5.31034 | 1.918386 | 0.111031 | 2.371559 | 1.613532 | 1.280956 | 1.124328 | -0.152 | 0.948601 | 0.687061 | 3.384741 | -2.32193 | -0.20091 | 1.925999 | 0.459432 | 0.67135 |
| HC01 | 1.499272 | 5.792335 | 5.009728 | 4.740658 | 7.456806 | 12.07709 | 1.166715 | 5.811728 | 5.1534 | 5.5627 | 5.755155 | 2.332565 | 2.526069 | 0.879706 | 2.744161 | 2.061776 | 1.969012 | 0.250962 | 0.948601 | 2.765535 | 2.035624 | -1.88897 | -0.37707 | -0.49411 | 0.063503 | 1.29071 |
| HC02 | 1.469886 | 4.853646 | 5.52979 | 6.508825 | 8.463054 | 14.34377 | 1.976364 | -0.074 | 6.65392 | 6.641979 | 5.755155 | 0.695994 | 2.526069 | 1.472488 | 4.490891 | -1.21759 | 1.684819 | 1.516015 | 0.077243 | 3.130107 | 2.134221 | -1.1016 | -0.37707 | 1.313246 | 0.286881 | 0.73372 |
| HC03 | 7.16115 | 8.00613 | 8.468747 | 7.812467 | 9.235054 | 12.90991 | 8.917217 | 8.092609 | 7.565396 | 4.180374 | 5.745093 | 4.952036 | 0.807489 | 0.97728 | 5.234904 | 2.783561 | 4.583158 | 3.021346 | 1.691981 | 4.155142 | 1.467279 | -1.2863 | 1.05641 | 2.886989 | 2.778629 | 2.40469 |
| HC04 | 5.192328 | 6.938332 | 7.42687 | 6.146722 | 9.04721 | 11.02825 | 7.353418 | 7.823812 | 7.46809 | 3.650788 | 3.653145 | 4.161742 | 0.285438 | 0 | 2.62293 | 1.495388 | 2.404549 | 2.64113 | -0.51457 | 4.045487 | 1.292782 | -1.2863 | 1.13606 | 2.617839 | 1.721023 | 0.71222 |
| SEP001 | 1.914565 | 5.658211 | 5.452974 | 4.868884 | 10.00253 | 9.689579 | 2.65764 | 1.9855 | 5.049413 | 5.299574 | 5.31034 | 1.918386 | 0.111031 | 1.217231 | 1.613532 | -0.30401 | 8.258001 | -0.152 | 3.403268 | 0.687061 | 0.687061 | -2.29336 | -0.21759 | 1.003602 | 3.674687 | 1.04985 |
| SEP002 | 3.532878 | 6.922049 | 2.74082 | 9.33558 | 8.601734 | 6.83706 | 2.65764 | 2.178237 | 1.361768 | 2.792855 | 5.31034 | 1.918386 | 2.169925 | -0.0145 | 1.613532 | -0.78588 | 7.958785 | -0.152 | 0.678072 | 0.687061 | 0.687061 | -2.32193 | -0.21759 | 0.823749 | 1.754888 | 2.77028 |
| SEP003 | 6.135053 | 10.90654 | 5.095924 | 6.584587 | 8.897588 | 10.08164 | 4.378512 | 7.567511 | 5.043301 | 5.90147 | 5.31034 | 5.83967 | 0.111031 | 1.99458 | 6.20183 | 3.731183 | 8.747438 | 0.719403 | 4.932864 | 3.912171 | 2.777157 | -2.30401 | 5.675393 | 3.740388 | 3.034744 | 2.87351 |
| SEP004 | 5.3067361 | 7.2931175 | 5.3708443 | 7.4376272 | 11.006603 | 12.957731 | 2.6576400 | 1.9855004 | 7.1763726 | 9.1955321 | 5.3103401 | 1.9183862 | 2.7420062 | 1.0036022 | 1.6135316 | 1.9278964 | 10.582184 | -0.152003 | 4.3150589 | 0.7136958 | 0.6870606 | -2.321928 | -0.2175914 | 1.7548875 | 2.4249220 | 2.07996 |
| SEP005 | 9.158925 | 6.584211 | 2.06695 | 5.120393 | 10.02302 | 12.41367 | -0.21759 | 8.949752 | 2.307866 | 7.911323 | 7.44931 | -2.36959 | 3.971452 | -0.05889 | -0.47393 | -0.76121 | 12.65734 | 3.061776 | 3.286881 | -0.23447 | -0.20091 | -2.07704 | -0.81097 | 1.22033 | 0.978196 | 1.92727 |
| SEP006 | -0.97143 | 6.624101 | 5.602291 | 5.431456 | 5.986638 | 11.30835 | -0.21759 | 8.113391 | -0.2034 | 7.154919 | -2.8365 | 0.309176 | -1.12186 | 1.111031 | -0.47393 | -1.53533 | 4.540089 | -0.41504 | 1.028569 | 0.378512 | -0.0145 | -0.87965 | 0.695994 | 1.831877 | -1.51457 | 1.62189 |
| SEP007 | 9.7806217 | 8.8441094 | 6.1576501 | 8.9724341 | 8.4536204 | 8.7842994 | 10.373262 | 10.024877 | 3.2128805 | 6.0851484 | 7.2627338 | 3.0452682 | 3.6898023 | 5.6064422 | 7.4134163 | 6.4016475 | 5.3300205 | 7.4144315 | 4.2326607 | 6.0624239 | 7.2076488 | 1.2418401 | 5.9551267 | 5.2966409 | 5.8398336 | 0.51437 |
| SEP008 | -0.9714306 | 6.9747587 | 4.8977244 | 3.5879649 | 7.1161359 | 10.115844 | -0.2175914 | 8.0238264 | -0.0854355 | -4.321928 | 1.6191782 | -1.4900509 | -2.3808218 | 0.6322682 | -0.4739312 | -1.3770696 | 3.8283269 | -0.4150375 | 0.4114262 | -0.2344653 | -0.2009127 | -1.962969 | -0.7490384 | 1.0840642 | -1.5145732 | 1.87996 |
| SEP009 | -0.9714306 | 9.1513590 | 6.2730495 | 5.9208887 | 10.395748 | 12.721772 | -0.2175914 | 9.4121845 | 0.0363274 | -4.321928 | 2.9893207 | -0.1648844 | 0.6798741 | -0.0588937 | -0.4739312 | -1.4540316 | 8.9999859 | -0.4150375 | 1.2418401 | 7.3675021 | -0.2009127 | -0.903738 | -0.7369656 | 1.7180875 | 4.1456774 | 1.52512 |
| SEP010 | -0.97143 | 7.149341 | 5.575312 | 6.030667 | 8.454731 | 13.02023 | -0.21759 | 8.812103 | 1.847596 | 2.980757 | 7.577176 | 0.152508 | 2.076217 | 0.163499 | -0.47393 | 1.003602 | 9.007462 | 1.632268 | 4.054848 | 3.643279 | 1.910733 | -1.56277 | 0.104337 | 1.731183 | 3.093391 | 1.21974 |
| SEP011 | -0.97143 | 4.556123 | 3.934988 | 5.606442 | 6.483413 | 10.5697 | -0.21759 | 6.533742 | -0.6827 | -4.32193 | -2.8365 | -1.66429 | -2.45799 | -0.05889 | -0.47393 | 1.735522 | 1.114367 | 2.82985 | -0.68966 | -0.23447 | 0.02148 | -1.63263 | 0.646163 | 1.472488 | 1.372952 | 1.42404 |
| SEP012 | 8.4791537 | 7.9709686 | 5.0172538 | 4.3809375 | 7.9627514 | 12.471346 | 11.190460 | 10.874731 | 10.627026 | 7.9274550 | 8.1216966 | 10.050107 | 9.9327159 | 8.5609059 | 8.9276298 | 5.0733918 | 5.4105785 | 1.0496307 | 8.8985409 | 11.665033 | 8.6027174 | 7.9510048 | 7.6101389 | 12.840212 | 3.0866139 | 1.40039 |
| SEP013 | 7.0230191 | 7.2566252 | 8.3727749 | 9.8108141 | 9.6399059 | 14.400038 | 7.9146350 | 7.8067449 | 8.3554828 | 4.34697* | 6.2978154 | 5.6823933 | 3.4661989 | 2.7297920 | 3.6171574 | 0.2329063 | 5.5228153 | 2.220949* | 0.5345596 | 4.7095663 | 1.6633446 | -1.2863042 | 2.4868430 | 2.2021355 | 2.8817429 | 2.43265 |
| SEP014 | 6.912576 | 6.618132 | 9.221738 | 7.604358 | 8.740543 | 12.92023 | 8.312439 | 7.916145 | 8.169064 | 5.16217 | 5.368058 | 6.496147 | 2.126765 | 0 | 2.62293 | 1.419755 | 4.875579 | 1.988776 | -0.51457 | 3.286881 | 1.443341 | -0.30847 | 0.570171 | 1.269033 | 3.036081 | 1.33372 |
| SEP015 | 5.513603 | 7.54591 | 8.953334 | 6.901081 | 9.564046 | 12.04345 | 7.33878 | 7.605516 | 6.447725 | 5.184708 | 4.342947 | 5.688449 | 1.38303 | 0 | 2.62293 | -0.91594 | 3.135272 | 1.6124 | -0.51457 | 4.626673 | 1.539531 | 0.859493 | 1.371392 | 3.557876 | 1.496922 | 1.12727 |
| SEP016 | 5.950702 | 6.204376 | 7.988741 | 7.438376 | 8.206233 | 11.29258 | 7.714108 | 7.916441 | 8.08192 | 5.204258 | 5.930524 | 6.187075 | 1.641084 | 0 | 4.221104 | 1.422233 | 4.675816 | 1.333424 | 2.02148 | 6.982537 | 3.883621 | 1.189034 | 2.765535 | 5.792335 | 2.650765 | 0.72297 |
| SEP017 | 11.75793 | 10.80485 | 8.457022 | 5.902978 | 14.07392 | 9.313223 | 8.581789 | 9.638036 | 8.589768 | 11.36062 | 9.161206 | 6.361187 | 8.809551 | 0 | 2.62293 | 1.584963 | 17.24386 | 1.314697 | 10.97604 | 8.251482 | 5.130313 | 3.380245 | 0.350497 | 1.269033 | 4.061776 | 0.35092 |
| SEP018 | 8.203935 | 6.364397 | 7.902797 | 7.245267 | 8.732473 | 9.777666 | 9.442259 | 8.1056 | 7.564203 | 6.543279 | 5.653805 | 7.885806 | 3.956243 | 3.877744 | 4.037382 | 5.160275 | 4.426265 | 5.39163 | 3.893362 | 9.289696 | 4.956057 | 3.82171 | 5.14323 | 7.543496 | 4.094236 | 0.34017 |
| SEP019 | 8.1003009 | 10.591855 | 6.0422065 | 5.4676055 | 9.6963264 | 12.047992 | 8.0545025 | 8.3822592 | 6.2789256 | 6.0336845 | 7.8046634 | 7.5501984 | 2.9713136 | 1.9541963 | 3.9097731 | 1.9212458 | 5.8939082 | 1.7708290 | 3.7114949 | 7.4350451 | 3.0916998 | 1.3715588 | 2.0045013 | 5.2331403 | 2.5607149 | 2.73157 |
| SEP020 | 6.016772 | 8.483693 | 7.666955 | 6.807216 | 9.090906 | 11.69311 | 6.894937 | 9.065662 | 6.542128 | 5.090045 | 6.235116 | 6.738734 | 3.418607 | 2.308885 | 2.62293 | 1.210856 | 7.360294 | 0.111031 | 5.05284 | 6.913126 | 1.981122 | 1.15866 | 1.654894 | 5.132803 | 1.376707 | 1.72297 |

| | 3- BCP panel (% of cells) | | | | | | | | | | | | | | | BCP type % with markers | | | | |
|---|---|---|---|---|---|---|---|---|---|---|---|---|---|---|---|---|---|---|---|---|
| Individuals | G CCR1+ | G CCR2+ | G CCR5+ | G CCR7+ | G TLR2+ | M CCR1+ | M CCR2+ | M CCR5+ | M CCR7+ | M TLR2+ | L CCR1+ | L CCR2+ | L CCR5+ | L CCR7+ | L TLR2+ | CD3 CCR1+ | CD3 CCR2+ | CD3 CCR5+ | CD3 CCR7+ | CD3 TLR2+ |
| Helathy donor 1 | 60.22 | 94.01 | 0.27 | 75.87 | 2.69 | 44.94 | 47.23 | 0.38 | 50.91 | 0.1 | 16.27 | 17.11 | 4.44 | 69.84 | 0.02 | 6.84 | 11.17 | 7.35 | 87.19 | 0.11 |
| HD280120553 | 0.17 | 19.53 | 0.5 | 15.89 | 24.21 | 12.13 | 87.02 | 5.62 | 81.6 | 84.37 | 1.29 | 21.79 | 2.62 | 15.58 | 7.4 | 4.42 | 3.49 | 2.34 | 74.48 | 25.98 |
| HD1202201280 | 1.32 | | 1.89 | 3.32 | 33.57 | 78.53 | | 5.3 | 80.3 | 64.3 | 8.69 | | 16.4 | 61.64 | 0.95 | 11.42 | | 14.17 | 73.53 | 0.19 |
| HD12020202273 | 0.73 | | 5.13 | 1.47 | 67.4 | 71.27 | | 3.24 | 64.8 | 72.72 | 26.71 | | 12.77 | 59.13 | 1.41 | 10.55 | | 12.75 | 77.93 | 0.23 |
| HD180220492 | 4.54 | 4.64 | 0.84 | 9.42 | 40 | 41.52 | 27.46 | 7.39 | 54.99 | 30.46 | 25.59 | 0.46 | 24.83 | 36 | 1.31 | 6.45 | 4.69 | 19.21 | 34.12 | 0.52 |
| HD030320137 | 2.72 | 7.25 | 5.14 | 2.66 | 51.36 | 62.02 | 62.14 | 13.63 | 85.57 | 59.85 | 4.59 | 12.49 | 3.7 | 56.21 | 2.56 | 6.88 | 14.12 | 1.17 | 62.48 | 0.6 |
| HD0503201179 | 1.53 | 43.72 | 2.04 | 23.17 | 89.52 | 48.18 | 53.54 | 13.35 | 74.11 | 58.64 | 4.48 | 7.97 | 9.49 | 69.82 | 3.26 | 5.23 | 6.85 | 7.51 | 79.48 | 0.26 |
| HD0503202152 | 2.44 | 2.61 | 2.96 | 8.89 | 1.22 | 61.1 | 64.73 | 8.68 | 83.96 | 57.91 | 6.61 | 10.54 | 14.57 | 65.43 | 1.36 | 5.43 | 10.47 | 10.59 | 77 | 0.23 |
| HC01 | 44.23 | 5.6 | 2.47 | 47.05 | 71.29 | 59.97 | 58.23 | 6.6 | 66.45 | 52.14 | 38.22 | 9.98 | 12.14 | 55.1 | 2.07 | 13.44 | 12.68 | 7.35 | 78.34 | 3.45 |
| HC02 | 56.43 | 10.23 | 0.48 | 29.18 | 57.76 | 36.8 | 33.34 | 3.28 | 75.76 | 33.29 | 21.5 | 9.14 | 5.1 | 70.48 | 0.87 | 2.61 | 11.57 | 6.08 | 86.6 | 0.23 |
| HC03 | 9.56 | 29.73 | 53.46 | 1.13 | 68.51 | 69.21 | 66.11 | 36.54 | 70 | 78.68 | 18 | 16.63 | 27.29 | 57.59 | 3.41 | 8.09 | 16.69 | 25.13 | 67.59 | 0.38 |
| HC04 | 11.35 | 35.4 | 32.22 | 1.51 | 39.33 | 63.51 | 69.58 | 55.8 | 59.48 | 67.02 | 10 | 18.86 | 19.38 | 66.65 | 1.28 | 2.08 | 18.54 | 15.28 | 77.67 | 0.14 |
| SEP001 | 82.59 | 3.49 | 0.81 | 14.9 | 19.16 | 74.26 | 63.43 | 1.59 | 69.63 | 69.3 | 6.68 | 4.09 | 6.3 | 39.72 | 1.54 | 8.17 | 7.67 | 3.92 | 69.09 | 0.34 |
| SEP002 | 1.4 | 0.54 | 0.56 | 9.51 | 15.24 | 72.36 | 70.12 | 4.74 | 72.7 | 79.51 | 8.05 | 4.71 | 4.36 | 67.14 | 2.68 | 5.17 | 4.78 | 5.97 | 89.35 | 0.17 |
| SEP003 | 65.55 | 6.91 | 3.01 | 25.33 | 28.93 | 80.29 | 35.66 | 25.05 | 81.9 | 59.71 | 17.54 | 8.25 | 25.69 | 53.91 | 2.97 | 22 | 3.35 | 24.91 | 50.4 | 2.41 |
| SEP004 | 49.96 | 0.96 | 0.04 | 35.6 | 54.26 | 96.56 | 81.67 | 1.16 | 94.29 | 95.29 | 40.44 | 7.65 | 2.16 | 58.59 | 5.74 | 7.82 | 7.45 | 1.31 | 75.36 | 1.42 |
| SEP005 | 7.09 | 1.68 | 0.04 | 9.79 | 13.47 | 68.6 | 59.68 | 0.62 | 56.3 | 66.34 | 24.61 | 1.3 | 4.52 | 29.64 | 0.92 | 13.5 | 3.52 | 15.03 | 75.41 | 0.41 |
| SEP006 | 16.5 | 50.61 | 0.59 | 10.77 | 52.58 | 71.24 | 70.12 | 2.18 | 70 | 74.54 | 12.57 | 7.86 | 11.69 | 52.38 | 2.1 | 7.14 | 6.13 | 14.25 | 56.51 | 2.42 |
| SEP007 | 10.33 | 44.19 | 4.47 | 5.67 | 26.17 | 76.96 | 76.02 | 6.02 | 82.07 | 90.26 | 12.1 | 23.62 | 13.22 | 37.02 | 2.35 | 5.47 | 22.24 | 20.08 | 72.63 | 3.48 |
| SEP008 | 2.52 | 6.69 | 3.78 | 14.53 | 6.1 | 3.5 | 3.17 | 1.35 | 27.5 | 8.67 | 30 | 0.67 | 1.99 | 41.57 | 1.4 | 5.4 | 4.77 | 13.65 | 57.7 | 6.88 |
| SEP009 | 41.67 | 37.5 | 0.2 | 22.85 | 29.25 | 47.43 | 44.32 | 2.59 | 55.29 | 68.63 | 8.9 | 17.99 | 7.1 | 69.67 | 11.56 | 4.03 | 10.06 | 6.03 | 92.63 | 3.74 |
| SEP010 | 7.27 | 13.78 | 0.53 | 9.07 | 9.83 | 88.28 | 80.67 | 5.73 | 90.46 | 90.42 | 12 | 23.95 | 10.1 | 46.1 | 5.25 | 8.8 | 16.41 | 13.5 | 76.7 | 7.24 |
| SEP011 | 15.6 | 64.75 | 3.01 | 37.63 | 28.69 | 71.18 | 68.86 | 8.39 | 94.37 | 93.61 | 7.11 | 18.15 | 22.3 | 54.97 | 4.61 | 2.36 | 12.43 | 14.43 | 57.67 | 1.82 |
| SEP012 | 12.5 | 36.25 | 3.42 | 41.84 | 33.88 | 88.97 | 85.1 | 6.09 | 96.78 | 89.93 | 8.56 | 23.08 | 25.63 | 78.33 | 4.73 | 3.81 | 16.64 | 17.74 | 80.94 | 2.08 |
| SEP013 | 7.24 | 30.92 | 1.32 | 22.88 | 20.89 | 15.51 | 12.54 | 1.31 | 32.23 | 14.36 | 5.93 | 5.1 | 3.57 | 79.61 | 0.91 | 4.87 | 3.59 | 2.36 | 95.9 | 0.55 |
| SEP014 | 10.72 | 41.33 | 0.08 | 17.66 | 59.4 | 58.54 | 63.47 | 2.37 | 73.86 | 62.26 | 3 | 4.93 | 4.29 | 30.19 | 0.77 | 3.58 | 18.72 | 15.57 | 85.42 | 2.65 |
| SEP015 | 19.21 | 33.72 | 0.96 | 17.4 | 13.02 | 65.57 | 68.16 | 8.23 | 76.2 | 68.9 | 3.12 | 10.52 | 27.97 | 30.81 | 2.1 | 2.13 | 11.2 | 18.65 | 34.76 | 2.24 |
| SEP016 | 8.08 | 27.01 | 0.12 | 8.8 | 35.78 | 43.62 | 49.46 | 0.55 | 60.75 | 51.24 | 7.48 | 4.43 | 8.05 | 55.99 | 3.71 | 2.73 | 5.58 | 9.6 | 77.07 | 2.99 |
| SEP017 | 15.54 | 0.25 | 0.05 | 11.83 | 0.56 | 3.18 | 1.44 | 0.44 | 30.12 | 3.19 | 8.71 | 4.87 | 12.32 | 39.72 | 1.58 | 8.63 | 2.4 | 22.1 | 50.54 | 3.01 |
| SEP018 | 3.79 | 3.92 | 1.38 | 9.97 | 6.02 | 47.05 | 44.66 | 2.65 | 52.2 | 48.15 | 4.76 | 7.02 | 6.52 | 33.09 | 1.46 | 18.04 | 15.11 | 16.76 | 81.11 | 8.97 |
| SEP019 | 2.82 | 6.07 | 0.95 | 10.81 | 1.41 | 57.89 | 41.94 | 23.52 | 69.2 | 51.46 | 9.38 | 9.57 | 7.85 | 72.58 | 1.82 | 6.28 | 7.73 | 9.53 | 83.62 | 1.51 |
| SEP020 | 0 | 52.38 | 17.08 | 38.73 | 54.16 | 86.96 | 83.14 | 7.6 | 94.69 | 94.9 | 12.78 | 23.37 | 18.86 | 42.71 | 7.84 | 5.24 | 18.3 | 33.39 | 49.79 | 2.58 |

[illegible]
